# Cardiovascular Diseases Burden in COVID-19: Systematic Review and Meta-analysis

**DOI:** 10.1101/2020.04.12.20062869

**Authors:** Amirhossein Hessami, Amir Shamshirian, Keyvan Heydari, Fatemeh Pourali, Reza Alizadeh-Navaei, Mahmood Moosazadeh, Saeed Abrotan, Layla Shojaei, Sogol Sedighi, Danial Shamshirian, Nima Rezaei

## Abstract

**Background:** High rate of cardiovascular disease (CVD) have been reported among patients with novel coronavirus disease (COVID-19). Meanwhile there were controversies among different studies about CVD burden in COVID-19 patients. Hence, we aimed to study CVD burden among COVID-19 patients, using a systematic review and meta-analysis.

**Methods:** We have systematically searched databases including PubMed, Embase, Cochrane Library, Scopus, Web of Science as well as medRxiv pre-print database. Hand searched was also conducted in journal websites and Google Scholar. Meta-analyses were carried out for Odds Ratio (OR) of mortality and Intensive Care Unit (ICU) admission for different CVDs. We have also performed a descriptive meta-analysis on different CVDs.

**Results:** Fifty-six studies entered into meta-analysis for ICU admission and mortality outcome and 198 papers for descriptive outcomes, including 159,698 COVID-19 patients. Results of meta-analysis indicated that acute cardiac injury, (OR: 13.29, 95% CI 7.35-24.03), hypertension (OR: 2.60, 95% CI 2.11-3.19), heart Failure (OR: 6.72, 95% CI 3.34-13.52), arrhythmia (OR: 2.75, 95% CI 1.43-5.25), coronary artery disease (OR: 3.78, 95% CI 2.42-5.90), and cardiovascular disease (OR: 2.61, 95% CI 1.89-3.62) were significantly associated with mortality. Arrhythmia (OR: 7.03, 95% CI 2.79-17.69), acute cardiac injury (OR: 15.58, 95% CI 5.15-47.12), coronary heart disease (OR: 2.61, 95% CI 1.09-6.26), cardiovascular disease (OR: 3.11, 95% CI 1.59-6.09), and hypertension (OR: 1.95, 95% CI 1.41-2.68) were also significantly associated with ICU admission in COVID-19 patients.

**Conclusion:** Findings of this study revealed a high burden of CVDs among COVID-19 patients, which was significantly associated with mortality and ICU admission. Proper management of CVD patients with COVID-19 and monitoring COVID-19 patients for acute cardiac conditions is highly recommended to prevent mortality and critical situations.

**Graphical abstract:** 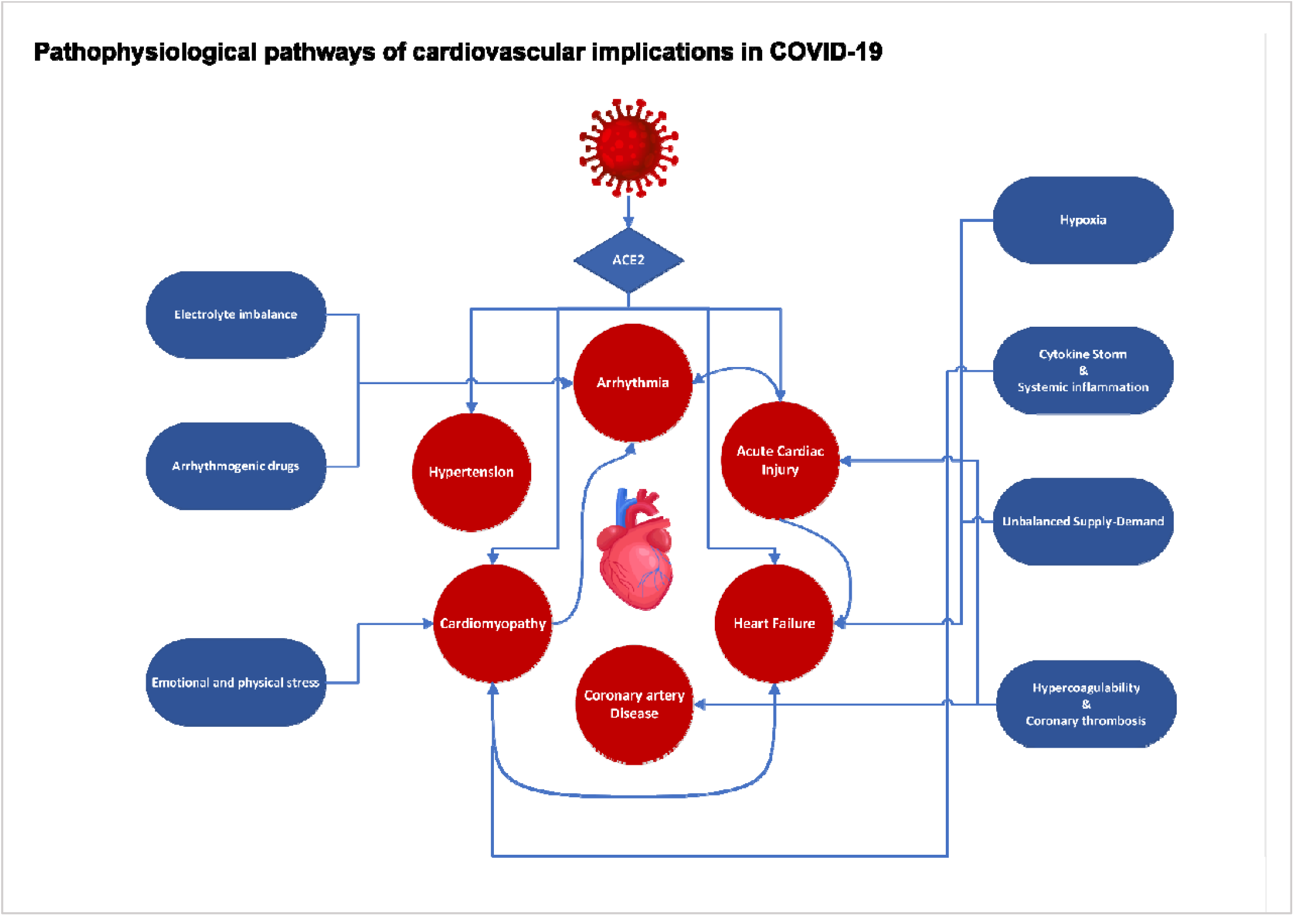

## Introduction

Coronaviruses are enveloped positive sense single stranded RNA viruses which cause respiratory infections in human and animals (1–3). There were six coronaviruses known to cause infection in humans (4), including Sever Acute Respiratory Syndrome (SARS), China, 2002 (5) and Middle Eastern Respiratory Syndrome (MERS), Saudi Arabia, 2012 (6, 7). The World Health Organization (WHO) reported cases of pneumonia with unknown source in Wuhan, China, December 2019 (8). Further investigations in samples of patients with respiratory infection, who were in contact with a seafood markets in Wuhan revealed a novel virus, named severe acute respiratory syndrome coronavirus 2 (SARS-CoV-2) (9). On 11^th^ March, 2020 WHO declared the SARS-CoV-2 outbreak as a pandemic (10). It has been estimated that 1.7 billion people (22% world population) have at least one underlying condition, including cardiovascular diseases (CVD), which increases the risk of developing severe disease in case of coronavirus disease (COVID-19) (11).

There is a mutual relationships between CVD and infections (12, 13), while the viral respiratory infectious diseases like influenza might increase the risk of myocardial infarction and cardiovascular events (13), underlying CVD might increase the risk of mortality among patients with infection (14).

Reports from COVID-19 disease including a large number of patients showed that fatality rate was 10.5% for CVD and 6.0% for hypertension among 72314 cases of COVID-19 (15). Studies indicated that there is an increased risk of mortality among hospitalized COVID-19 patients due to CVD (16–18).

Given the increasing number of COVID-19 patients besides from common clinical presentations of disease, CVD in COVID-19 infected patients are seems to be concerning (16). We aimed to assess the CVD burden of COVID-19, using a systematic review and meta-analysis method in this study.

## Methods

### Search Strategy

In this study, Preferred Reporting Items for Systematic Reviews and Meta-analyses (PRISMA) guideline was used for study design, search strategy, screening and reporting. The research question has been developed using PECO; “P” stands for Patients, “E” as Exposure, “C” as Comparison and “O” as Outcome. PECO components were as follows: “P”; hospitalized COVID-19 patients, “E”; CVDs, “C”; no CVD, “O”; ICU admission/mortality. A systematic search was using all available MeSH terms and free keywords for “COVID-19”, “Cardiovascular Disease” “Myocardial Infarction”, “Heart Failure”, “Hypertension”, “Myocarditis”, “Arrhythmia”. Searched Databases included PubMed, Embase, Scopus, Web of Science, Cochrane Library, medRxiv pre-print database as well as Science Direct search engine. Hand search was done in publishers and journals databases including: Center for Disease Control and Prevention (CDC), The Journal of the American Medical Association (JAMA), The Lancet, The British Medical Journal (BMJ), Nature, Wiley, New England Journal of Medicine, Cambridge and Oxford. Our search included papers in all languages and there was no time limitation for publications. All original Cohort, Case Control, Cross-Sectional and Case-Series studies until 27 ^th^ May 2020 were included.

### Criteria for Study Selection

Two members of our team (F.P and A.H) selected the study independently and in case of disagreement R.A made the final decision. Studies met the following criteria included into systematic review: 1) Studies reporting characteristics of hospitalized COVID-19 patients; 2) Studies which reported any CVD in COVID-19 patients; 3) COVID-19 confirmed by Chest CT Scan, RT-PCR and hallmarks of the disease. Criteria for including studies into meta-analysis were: 1) Studies that reported CVD in COVID-19 patients admitted to ICU; 2) Studies reported the mortality rate of COVID-19 patients with underlying CVD; 3) Studies reported CVD among COVID-19 hospitalized patients. Studies were excluded if they: 1) Reported outpatients or asymptomatic COVID-19 patients; 2) Not reported CVDs; 3) Review papers, case reports, *in vitro* studies and animal studies.

### Data Extraction

Two investigators (F.P and A.H) have independently evaluated quality of publications and extracted data from included papers. In case of disagreement a supervisor (R.A) solved the issue and made the final decision. Data extraction included first author name, publication year, country and following data extracted for each group (Total Sample, ICU, Non-ICU, Mortality, Survival): Sample size, mean ± standard deviation (SD) of age, number of females, number of males, heart failure, hypertension, other cardiovascular disease, acute cardiac injury, cardiomyopathy, myocardial damage, heart palpitation, coronary heart disease, arrhythmia and acute cardiac injury. In cases that data was presented as median (interquartile range), a method by *Wan et al*. (19) was used to calculate mean ± SD.

### Risk of Bias Assessment

Newcastle-Ottawa Scale tool was used for risk of bias assessment of studies included into meta-analysis (20). Risk of bias only assessed for studies entered into meta-analysis main outcomes (ICU and mortality) of the study (Figure 1).

**Figure 1.**
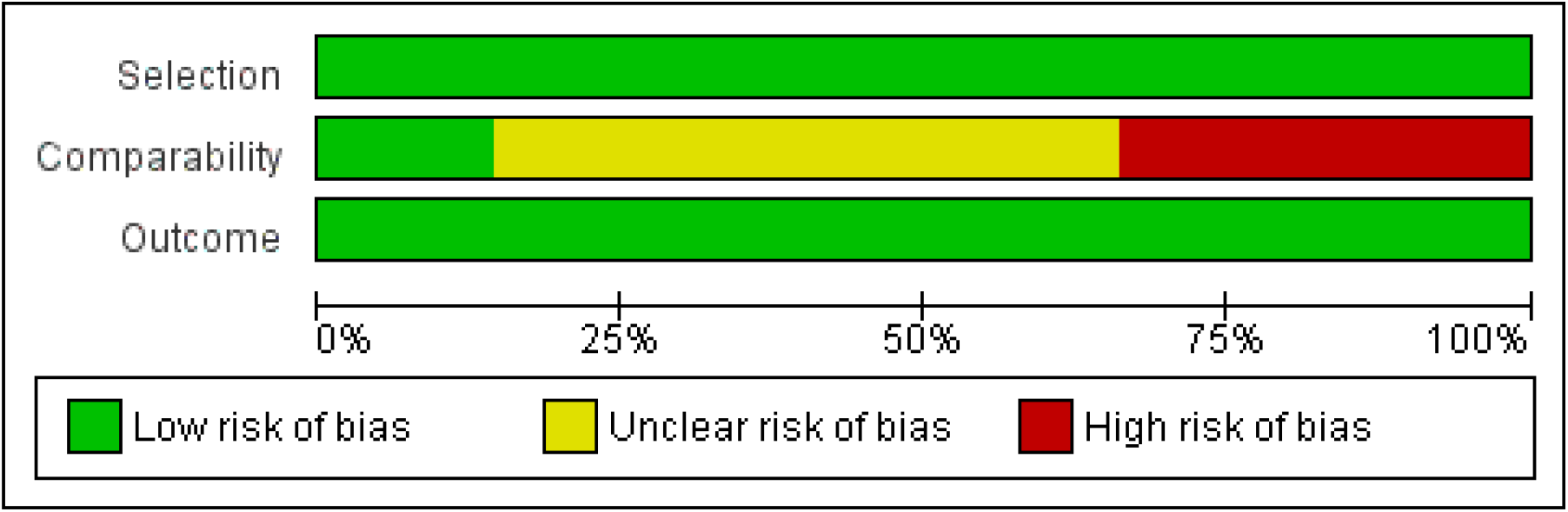
Summary of Risk of Bias

**Figure 1.**
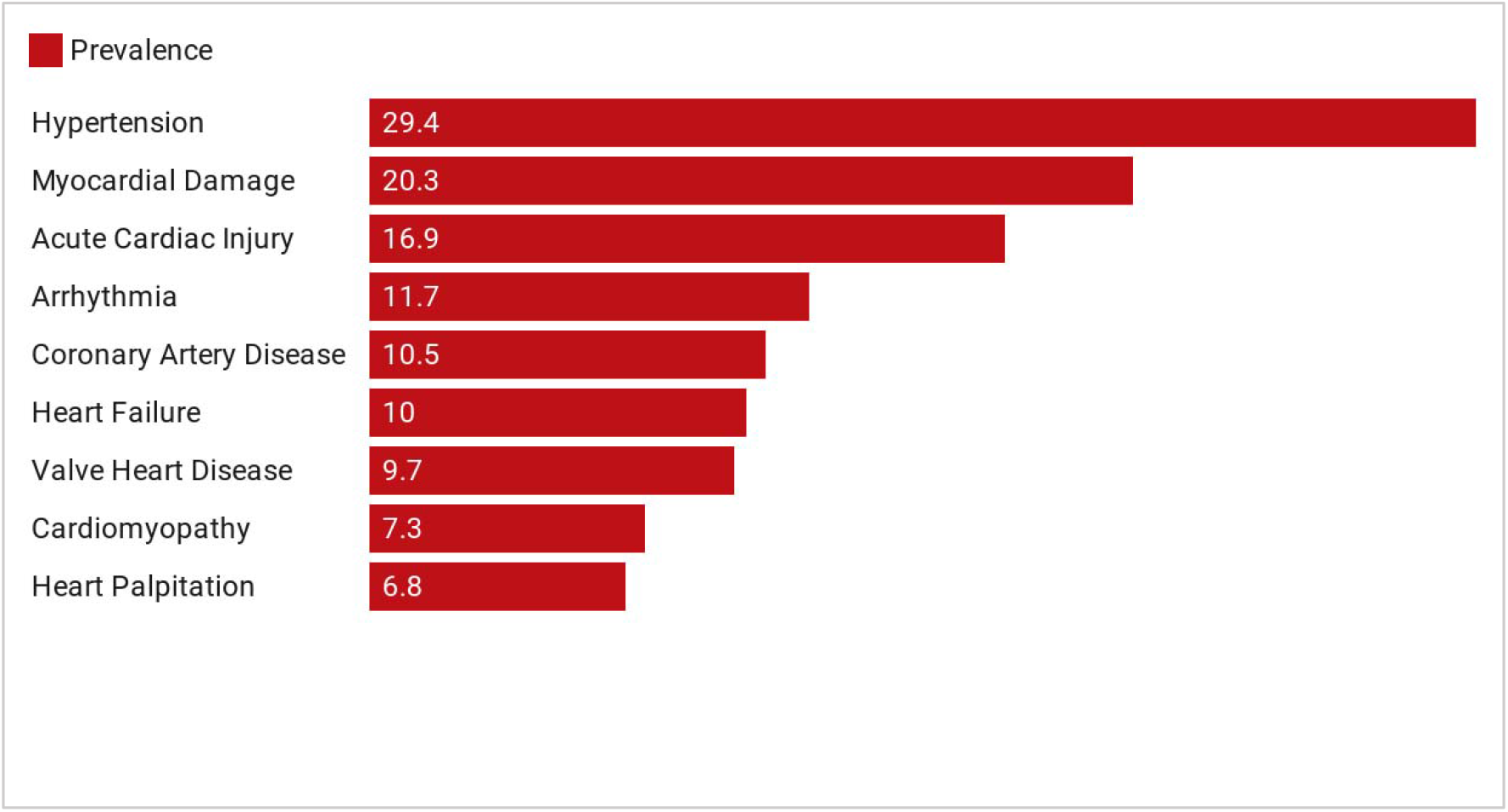
Prevalence of Cardiovascular Complications Among COVID-19 Patients

### Data Analysis

Odds Ratio (OR) and pooled estimate prevalence rate with 95% confidence interval (CI), were calculated using statistical analysis Comprehensive Meta-Analysis (CMA) V.2. In order to assess the heterogeneity, *I*-square (*I*^2^) test was used. In case of high heterogeneity (more than 50%) random effect model was used for meta-analysis. Publication bias has been assessed using *Begg*’s test.

## Results

### Study selection process

Our search through databases resulted in 4658 records. Duplicate records, including 1695 studies, have been excluded and after title and abstract screening, full texts of 2963 papers were assessed for eligibility. Four studies were excluded due to unavailability of the full-text. Finally, 56 papers (21–76) entered into meta-analysis for primary outcomes (mortality and ICU admission) and 198 papers (16, 21–217) for descriptive outcomes. PRISMA flow diagram for the study selection process is presented in Figure 2.

**Figure 2.**
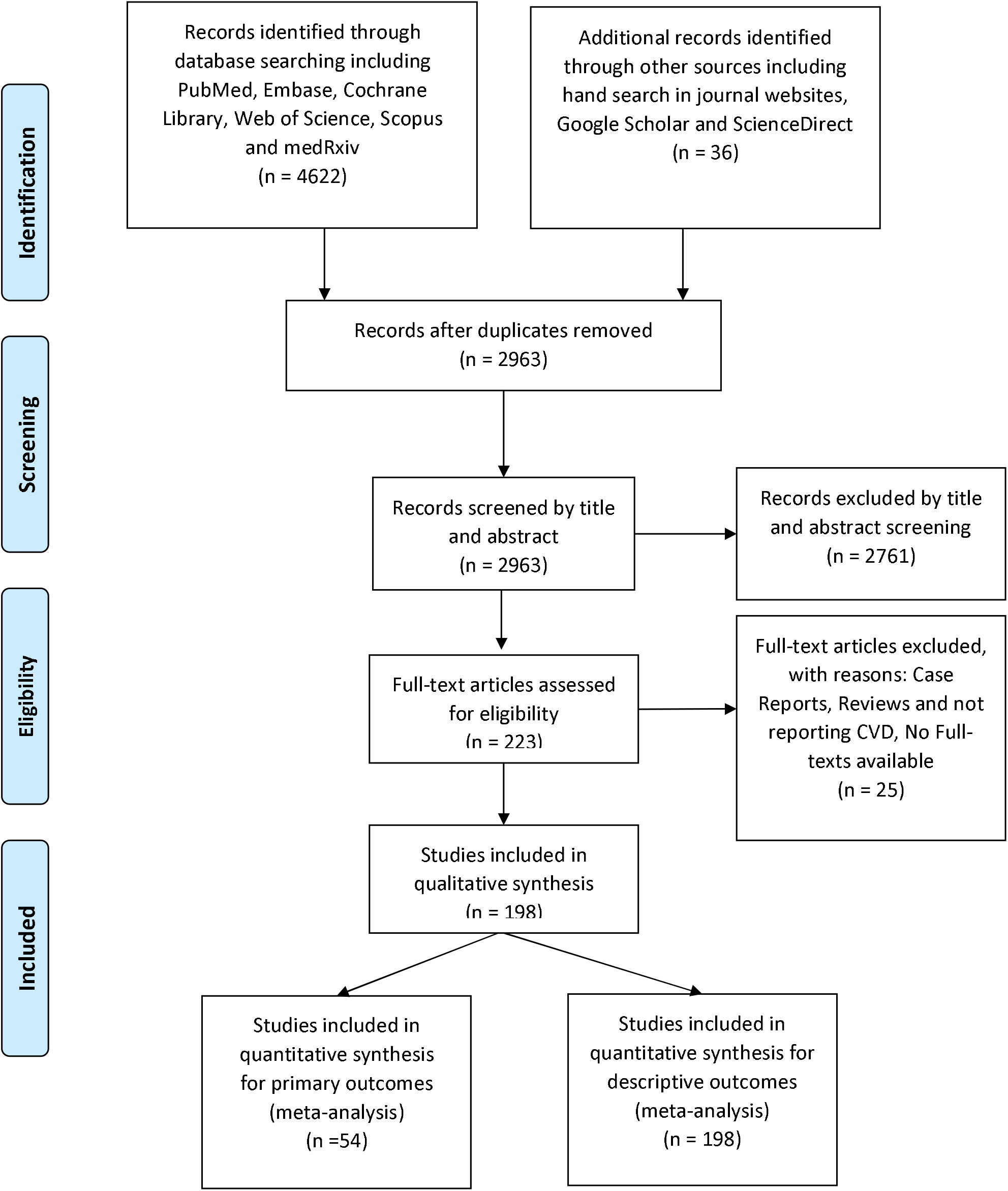
PRISMA Flow Diagram

### Study characteristics

Out of 56 papers included into meta-analysis for primary outcomes, 2 studies were case-control, 1 case-cohort, 8 case series and 45 cohorts. The studies’ sample size ranged from 13 to 11095 including 29,056 participants. One-hundred ninety-nine papers entered into meta-analysis for descriptive outcomes including 168 cohorts, 4 case-control, 4 cross-sectional, 21 case-series, 1 case-cohort. Studies sample size ranged from 10 to 27584 including 159698 patients.

### Quality assessment

According to NOS tool for quality assessment, 56 studies earned the minimum eligibility score and entered into the meta-analysis for primary outcomes. Summary of risk of bias is presented in Figure 2. *Begg*’s test showed that there was no publication bias (*P*=0.7).

### Mortality

The meta-analysis showed that prevalence of CVDs among COVID-19 patients with mortality were as follows: Acute Cardiac Injury (52%), Hypertension (51%), Arrhythmia (37%), Heart Failure (27%), Coronary Heart Disease (23%) and Cardiovascular Diseases (23%) (Table 1).

**Table 1.**
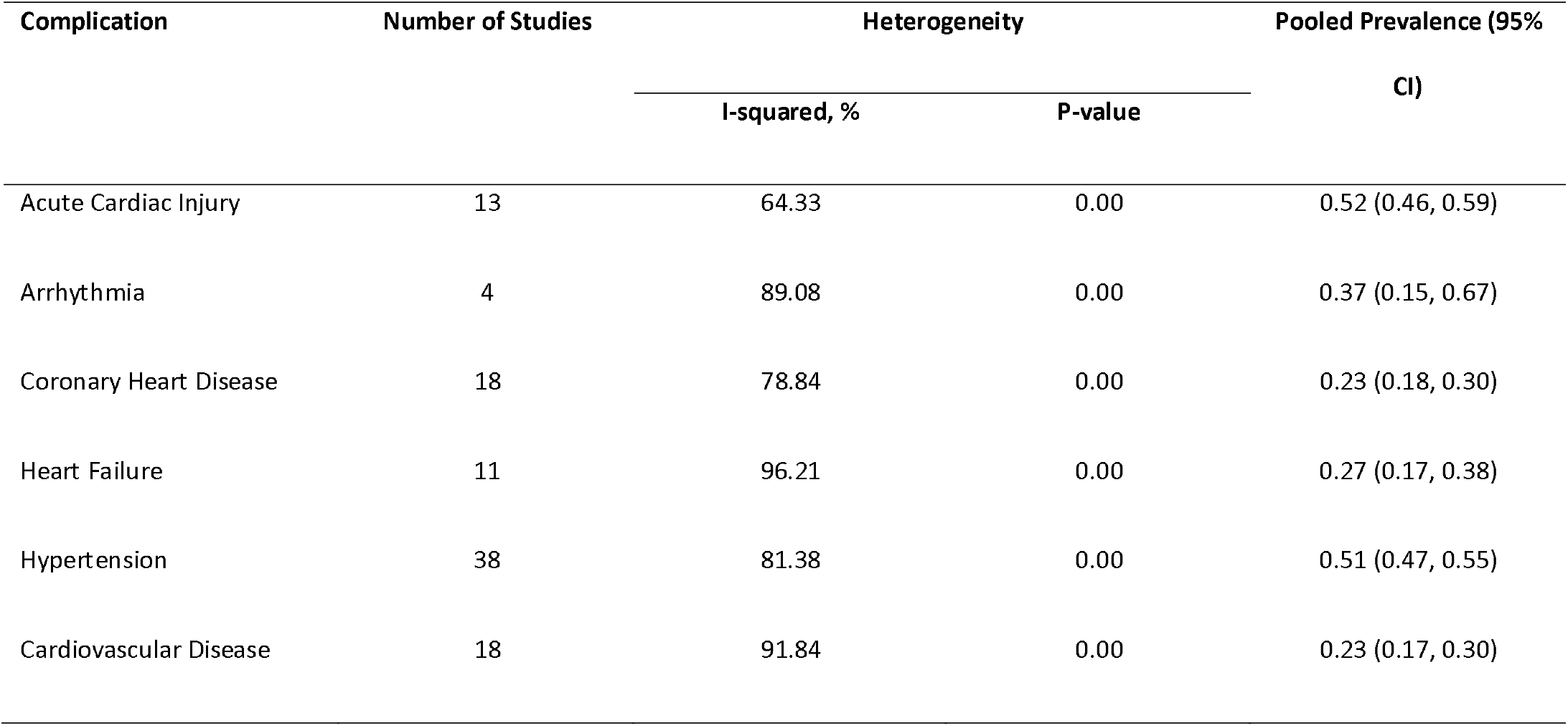
Meta-analysis of pooled estimate prevalence of CVDs among COVID-19 patients with mortality

Results of meta-analysis indicated that presence of Acute Cardiac Injury (OR: 13.29, 95% CI 7.35-24.03), Coronary Artery Disease (OR: 3.78 95% CI 2.42-5.90), Arrhythmia (OR: 2.75, 95% CI 1.43-5.25), Hypertension (OR: 2.60, 95% CI 2.11-3.19), Heart Failure (OR: 6.72, 95% CI 3.34-13.52), and Cardiovascular Disease (OR: 2.61, 95% CI 1.89-3.62) were significantly associated with mortality in COVID-19 patients (Table 2).

**Table 2.**
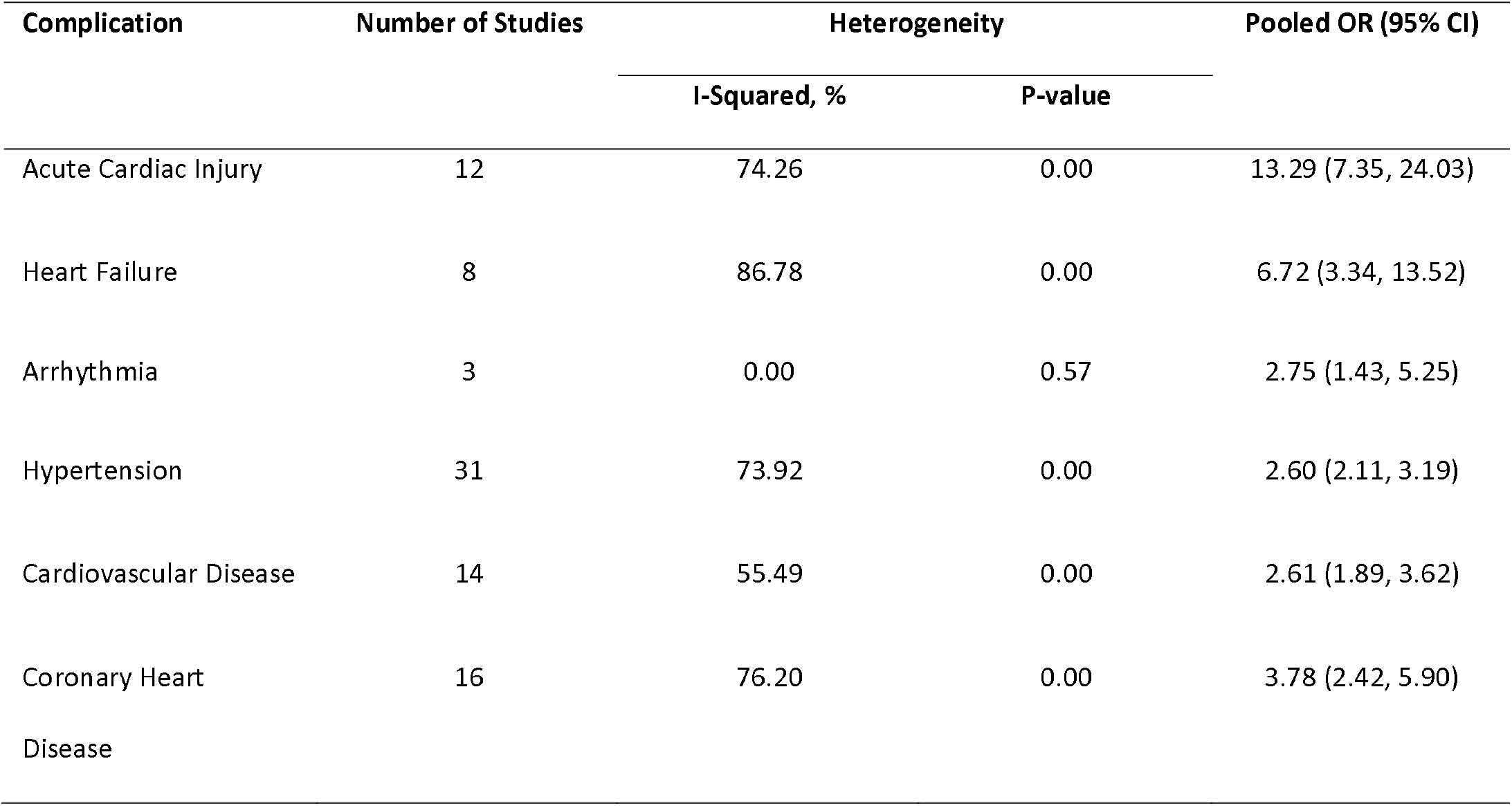
Meta-analysis of OR of mortality in COVID-19 patients for CVDs

| Complication | Number of Studies | Heterogeneity |  | Pooled OR (95% CI) |
| --- | --- | --- | --- | --- |
|  |  | I-Squared, % | P-value |  |
| Acute Cardiac Injury | 12 | 74.26 | 0.00 | 13.29 (7.35, 24.03) |
| Heart Failure | 8 | 86.78 | 0.00 | 6.72 (3.34, 13.52) |
| Arrhythmia | 3 | 0.00 | 0.57 | 2.75 (1.43, 5.25) |
| Hypertension | 31 | 73.92 | 0.00 | 2.60 (2.11, 3.19) |
| Cardiovascular Disease | 14 | 55.49 | 0.00 | 2.61 (1.89, 3.62) |
| Coronary Heart Disease | 16 | 76.20 | 0.00 | 3.78 (2.42, 5.90) |

### Intensive care unit admission

Pooled prevalence of cardiovascular implications including Acute Cardiac Injury, Arrhythmia, Heart Failure, Coronary Artery Disease, Hypertension and Cardiovascular Diseases were 33.6%, 33.0%, 20.4%, 20.6%, 43.6% and 25.0% in COVID-19 patients admitted to ICU respectively (Table 3).

**Table 3.** Meta-analysis of pooled estimate prevalence of CVDs among COVID-19 patients admitted to ICU

| Complication | Number of Studies | Heterogeneity |  | Pooled Prevalence (95% CI) |
| --- | --- | --- | --- | --- |
|  |  | I-squared, % | P-value |  |
| Acute Cardiac Injury | 8 | 51.15 | 0.04 | 0.33 (0.24, 0.43) |
| Arrhythmia | 3 | 71.88 | 0.02 | 0.33 (0.18, 0.51) |
| Coronary Heart Disease | 13 | 67.77 | 0.00 | 0.20 (0.15, 0.27) |
| Heart Failure | 7 | 78.42 | 0.00 | 0.20 (0.09, 0.37) |
| Hypertension | 31 | 88.89 | 0.00 | 0.43 (0.37, 0.50) |
| Cardiovascular Disease | 19 | 91.54 | 0.00 | 0.25 (0.16, 0.35) |

The meta-analysis on ICU outcome showed that the odds of ICU admission in COVID-19 patients is significantly associated with Acute Cardiac Injury (OR: 15.58, 95% CI 5.15 − 47.12), Arrhythmia (OR: 7.03, 95% CI 2.79 − 17.69), Coronary Heart Disease (OR: 2.61, 95% CI 1.09 − 6.26), Cardiovascular Disease (OR: 3.11, 95% CI 1.59 − 6.09) and Hypertension (OR: 1.95, 95% CI 1.41 − 2.68). Heart Failure was not statistically significantly (OR: 2.44, 95% CI 0.67 − 8.79) associated with ICU admission in COVID-19 however, the effect size was considerable (Table 4).

**Table 4.** Meta-analysis of OR for ICU admission outcome among COVID-19 patients

| Complication | Number of studies | Heterogeneity |  | Prevalence (95% CI) |
| --- | --- | --- | --- | --- |
|  |  | I-Square (%) | P-value |  |
| Coronary heart disease | 60 | 93.92 | 0.00 | 0.10 (0.09, 0.12) |
| Cardiovascular and cerebrovascular disease | 16 | 94.97 | 0.00 | 0.14 (0.09, 0.22) |
| Cardiovascular disease | 87 | 95.73 | 0.00 | 0.11 (0.09, 0.12) |
| Arrhythmia | 17 | 91.17 | 0.00 | 0.11 (0.07, 0.16) |
| Acute cardiac injury | 28 | 92.35 | 0.00 | 0.16 (0.13, 0.19) |
| Cardiomyopathy | 4 | 85.25 | 0.00 | 0.07 (0.01, 0.27) |
| Heart failure | 31 | 99.11 | 0.00 | 0.10 (0.07, 0.13) |
| Myocardial damage | 3 | 94.49 | 0.00 | 0.20 (0.06, 0.49) |
| Hypertension | 158 | 97.97 | 0.00 | 0.29 (0.27, 0.31) |
| Heart palpation | 6 | 85.06 | 0.00 | 0.06 (0.03, 0.13) |
| Heart Valve Disease | 3 | 87.00 | 0.00 | 0.09 (0.02, 0.36) |

### Cardiovascular Complications

One-hundred ninety-eight studies have investigated for cardiovascular complications in the COVID-19 patients. Pooled prevalence of cardiovascular complications observed among patients with COVID-19 included hypertension (29%), cardiovascular and cerebrovascular disease (14%), heart failure (10%), acute cardiac injury (16%), coronary heart disease (10%), myocardial damage (20%), cardiovascular disease (11%), arrhythmia (11%), cardiomyopathy (7%), heart palpitation (6%) and heart valve disease (9%) (Table 5, Figure 3).

**Table 5.** Pooled prevalence for cardiovascular complications among COVID-19 patients

| Complication | Number of Studies | Heterogeneity |  | Pooled OR (CI 95%) |
| --- | --- | --- | --- | --- |
|  |  | I-squared, % | P-value |  |
| Acute Cardiac Injury | 6 | 61.73 | 0.02 | 15.58 (5.15, 47.12) |
| Arrhythmia | 2 | 32.22 | 0.22 | 7.03 (2.79, 17.69) |
| Coronary Heart Disease | 8 | 77.65 | 0.00 | 2.61 (1.09, 6.26) |
| Hypertension | 21 | 67.62 | 0.00 | 1.95 (1.41, 2.68) |
| Cardiovascular Diseases | 12 | 71.01 | 0.00 | 3.11 (1.59, 6.09) |

We have also analyzed pooled prevalence of cardiovascular disease in COVID-19 patient in different countries. Results of meta-analysis were as follows: Brazil (50.0%), China (7.8%), France (48.0%), Germany (45.5%), Iran (4.4%), Italy (24.7%), Netherlands (44.0%), South Korea (11.2%), Spain (16.9%), Switzerland (71.4%), United Kingdome (15.1%), and United States (24.4%) (Figure 4).

**Figure 4.**
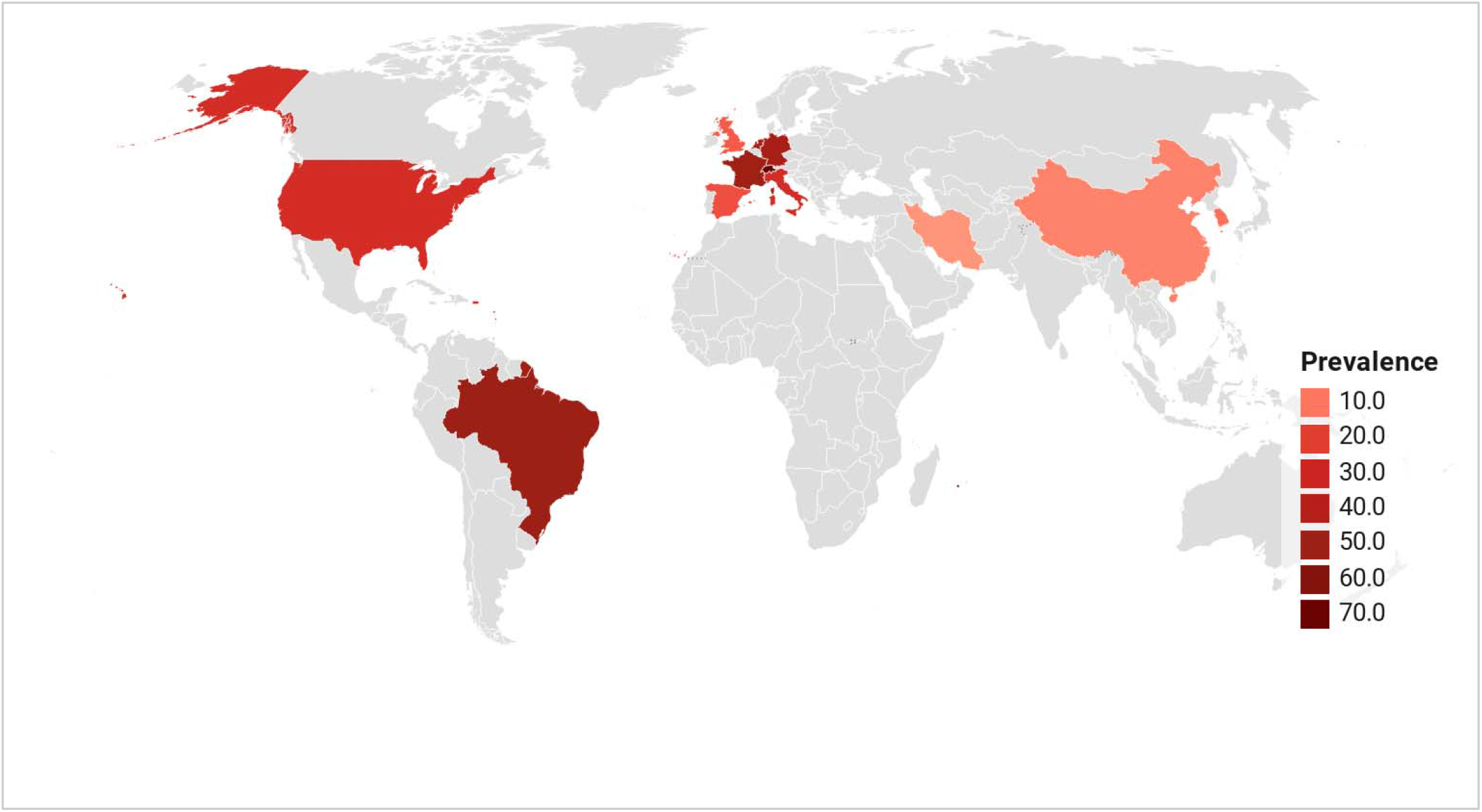
Cardiovascular Disease Burden among COVID-19 Patients in different countries

## Discussion

The results of this meta-analysis indicated cardiovascular implications including acute cardiac injury, arrhythmia, coronary heart disease, hypertension and cardiovascular diseases were significantly associated with COVID-19 patient’s admission to the ICU. Comparing pooled estimate of OR for CVDs showed odds of ICU admission was significantly higher in acute cardiac injury and arrhythmia than hypertension, but there was no significant difference between other cardiovascular implications. Investigations have shown that mortality in patients with acute cardiac injury was significantly higher than coronary artery disease, arrhythmia, and hypertension. Comparing estimated frequency of different cardiovascular complications including acute cardiac injury, arrhythmia, cardiomyopathy, coronary heart disease, heart palpitation, hypertension, myocardial damage, heart failure and other cardiovascular diseases did not show any significant difference among them.

Cardiovascular complications have been previously reported in previous respiratory infections with similar etiology and their condition affects severity of the disease (17, 218); so that even hospitalization for pneumonia is associated with long-term and short-term risk of CVD (12). Viral infections cause imbalance between cardiac supply and demand and increase in systemic inflammation. Therefore patients with pre-existing CVD have higher risks for acute cardiac conditions (219), infection, and develop severe conditions during the infection (220).

Acute myocardial injury was significantly associated with both ICU admission and mortality. There were several etiologies proposed for acute myocardial injury in these patients. The first possible mechanism is myocardial injury caused by cytokine storm as the results of systematic inflammation mediated by pathologic T-cells (221–223). Study by Chen *et al*. (25) showed elevated myocardial injury biomarkers including NT-proBNP, cTnI and hs-CRP were significantly correlated with COVID-19 severity. The second possible explanation is the imbalance between supply and demand caused by systemic infection along with hypoxia caused by respiratory infection, which lead to acute myocardial injury (221, 223). There are evidences that shows SARS-CoV-2 binds to human angiotensin converting enzyme-2 (ACE2) to infect the cells (224), which being highly expressed in lungs and heart. The binding of SARS-CoV-2 to ACE2 in heart can results acute myocardial injury (221, 225). COVID-19 has been associated with thrombotic events and coagulations disorders (226), which might lead to hypercoagulability and coronary thrombosis results in acute myocardial infarction (221, 223).

SARS-CoV-2 binding to ACE2 causes activation of renin-angiotensin system and its complications including hypertension, heart failure, and atherosclerosis (220, 227, 228) as we resulted in our meta-analysis. These data could also suggest a reason for high prevalence of hypertension in our pooled estimate. Underlying CVDs like hypertension could higher the risk of infection and developing more severe symptoms in COVID-19 patients (229). We showed in our meta-analysis that hypertension was significantly associated with ICU admission and mortality; therefore blood pressure control could be potentially beneficial to reduce disease burden (230). Using Angiotensin Receptor Blockers (ARBs) and ACE inhibitors in order to control hypertension have raised question in the era of the COVID-19 pandemic. Studies suggested that using ARBs and ACE inhibitors could potentially be harmful as they can cause up-regulation of ACE2, SARS-CoV-2 receptor, and lung injury (230, 231); so, they should be carefully considered in COVID-19 patients (229). This meta-analysis also indicated that mean age of COVID-19 patients admitted to ICU was significantly higher that non-ICU. An explanation to this condition by AlGhatrif *et al*. (232) suggested that elderly patients with hypertension are more likely to have down-regulation of ACE2 expression, due to viral binding, and up-regulated angiotensin II, which exaggerates pro-inflammatory condition, predisposing them to severe conditions and mortality (233).

Arrhythmia was significantly associated with ICU admission and mortality in our meta-analysis. There are pathophysiological mechanisms suggested for arrhythmia among COVID-19 patients; however, whether these patients had pre-existing arrhythmia or secondary to COVID-19 remains unknown (234). First, as any systematic infection COVID-19 can cause electrolyte imbalance, especially hypokalemia (235), which might contribute to arrhythmia in susceptible patients (221). Second, as we have discussed earlier myocardial injury has been an important subject in COVID-19 patients. It has been suggested that myocardial damage can be potentially triggers arrhythmia (236, 237). Third, pharmacological therapies prescribed for these patients can be potential risk factors for arrhythmia. Hydroxychloroquine, as one the most common drugs which had been used in COVID-19, can triggers QT prolongation (237, 238). Fourth, arrhythmia could also occur secondary to myocarditis by disrupting cardiomyocyte membrane and electrolyte imbalance, myocardial fibrosis, proinflammatory cytokines and edema of pericardium (234).

In our meta-analysis the prevalence of heart palpitation was relatively high and it has been reported as one the initial symptoms of the disease (239). National Health Commission of China (NHC) reported that some patients have presented with heart palpitations and chest tightness instead of cough and fever as initial symptoms of COVID-19 (229); so, heart palpitations in outpatients setting needs to carefully considered as potential symptom of COVID-19.

Heart failure was significantly associated with mortality in our meta-analysis, ranked after acute cardiac injury. The incidence of heart failure might be as the consequence of myocardial injury or myocarditis (240), which has been previously discussed. Increasing systemic metabolic demand as the results of systemic infection could also exacerbate previous stable heart failure (221). Right heart failure and pulmonary hypertension should be also carefully considered in patients with acute respiratory distress syndrome (ARDS) and parenchymal lung disease (236).

Cardiomyopathy has been also observed in COVID-19 patients in various forms. Takotsubo syndrome (TTS) or stress cardiomyopathy, characterized by transient left ventricular dysfunction, is triggered by emotional and physical stresses following to natural disasters (241), like COVID-19 pandemic. There were number of studies reported TTS in COVID-19 patients (242–246). Mechanism of stress cardiomyopathy in COVID-19 patients might be due to emotional distress caused by pandemic along with inflammatory and metabolic distress (244). Inflammatory cardiomyopathy or myocarditis was previously described in viral infections (247) as well as COVID-19 (248, 249). Cardiomyocytes expresses ACE2 which is the receptor for SARS-CoV-2, so this virus could infects human heart which can even exacerbate in case of heart failure (234).

Coronary artery disease was significantly associated with mortality of COVID-19 patients in our study. In patients with previous coronary artery disease, inflammatory state and hemodynamic changes in infection can potentially increase the risk of plaque rupture (163, 236, 250). Coagulopathy in COVID-19 patients (226) can also cause acute coronary syndrome in these patients.

We have discussed cardiovascular implications among COVID-19 patients. Cardiovascular diseases have significant role in the outcome of COVID-19 patients. Therefore, careful consideration and management of cardiovascular disease, from diagnosis to bedside, among COVID-19 patients are necessary. Results of this study could help policy makers, physicians and healthcare workers in front line to make evidence-based decisions and reduce the mortality and morbidity of this 21 st century pandemic.

Limitation of this study was high heterogeneity of studies in population. Compounding effects of other co-morbidities in ICU admission and mortality was not being considered. It is possible that other co-morbidities related to respiratory system, renal system and gastrointestinal system affects patient’s condition. Cardiovascular implications could be pre-existing in patients or either developed by the infection; so, we could not determine the casual relationship. However, given the burden and vital role of CVDs the importance does not differ. We only included studies of hospitalized adult COVID-19 patients and asymptomatic and outpatients are excluded. Data regarding to cardiovascular diseases in COVID-19 patients in some countries were missing.

## Conclusion

In conclusion, CVD have a significant role in disease severity and mortality of COVID-19 patients. Hypertension, acute cardiac injury, coronary heart diseases in COVID-19 patient needs to be carefully monitored and managed in case of acute conditions. Other cardiovascular implications, including arrhythmia and heart failure also need to be considered since they can be fatal. Therefore, careful consideration and management of cardiovascular disease among these patients are necessary. Results of this study could help policy makers, physicians and healthcare workers in front line to make evidence-based decisions and reduce the mortality and morbidity of this 21^st^ century pandemic.

## Data Availability

The data that support the findings of this study are openly available in data bases mentioned in the search strategy.

## Supplementary Material

Forest Plots and characteristics of studies included into meta-analyses are presented in supplementary material.

## Acknowledgments

We acknowledge Student Research Committee of Mazandaran University of Medical Sciences for supporting this project (code number: 7396).

## References

1. Cheng VCC, Lau SKP, Woo PCY, Yuen KY. Severe acute respiratory syndrome coronavirus as an agent of emerging and reemerging infection. Clin Microbiol Rev. 2007;20(4):660–94.

2. Lu R, Zhao X, Li J, Niu P, Yang B, Wu H, et al. Genomic characterisation and epidemiology of 2019 novel coronavirus: implications for virus origins and receptor binding. The Lancet. 2020;395(10224):565–74.

3. Saghazadeh A, Rezaei N. Immune-epidemiological parameters of the novel coronavirus -a perspective. Expert Rev Clin Immunol. 2020;16(5):465–70.

4. Su S, Wong G, Shi W, Liu J, Lai ACK, Zhou J, et al. Epidemiology, Genetic Recombination, and Pathogenesis of Coronaviruses. Trends in Microbiology. 2016;24(6):490–502.

5. Peiris J, Guan Y, Yuen K. Severe acute respiratory syndrome. Nature medicine. 2004;10(12):S88–S97.

6. Zaki AM, Van Boheemen S, Bestebroer TM, Osterhaus AD, Fouchier RA. Isolation of a novel coronavirus from a man with pneumonia in Saudi Arabia. New England Journal of Medicine. 2012;367(19):1814–20.

7. Lotfi M, Hamblin MR, Rezaei N. COVID-19: Transmission, prevention, and potential therapeutic opportunities. Clin Chim Acta. 2020;508:254–66.

8. Organization WH. Pneumonia of unknown cause–China. Emergencies preparedness, response, Disease outbreak news, World Health Organization (WHO). 2020.

9. Zhu N, Zhang D, Wang W, Li X, Yang B, Song J, et al. A novel coronavirus from patients with pneumonia in China, 2019. New England Journal of Medicine. 2020.

10. Organization WH. WHO Director-General’s opening remarks at the media briefing on COVID-19-11 March 2020. Geneva, Switzerland. 2020.

11. Clark A, Jit M, Warren-Gash C, Guthrie B, Wang HHX, Mercer SW, et al. Global, regional, and national estimates of the population at increased risk of severe COVID-19 due to underlying health conditions in 2020: a modelling study. The Lancet Global Health.

12. Corrales-Medina VF, Alvarez KN, Weissfeld LA, Angus DC, Chirinos JA, Chang C-CH, et al. Association between hospitalization for pneumonia and subsequent risk of cardiovascular disease. JAMA. 2015;313(3):264–74.

13. Madjid M, Miller CC, Zarubaev VV, Marinich IG, Kiselev OI, Lobzin YV, et al. Influenza epidemics and acute respiratory disease activity are associated with a surge in autopsy-confirmed coronary heart disease death: results from 8 years of autopsies in 34,892 subjects. Eur Heart J. 2007;28(10):1205–10.

14. Dhainaut J-F, Claessens Y-E, Janes J, Nelson DR. Underlying Disorders and Their Impact on the Host Response to Infection. Clinical Infectious Diseases. 2005;41(Supplement_7):S481–S9.

15. Wu Z, McGoogan JM. Characteristics of and Important Lessons From the Coronavirus Disease 2019 (COVID-19) Outbreak in China: Summary of a Report of 721314 Cases From the Chinese Center for Disease Control and Prevention. JAMA. 2020.

16. Guo T, Fan Y, Chen M, Wu X, Zhang L, He T, et al. Cardiovascular implications of fatal outcomes of patients with coronavirus disease 2019 (COVID-19). JAMA cardiology. 2020.

17. Madjid M, Safavi-Naeini P, Solomon SD, Vardeny O. Potential Effects of Coronaviruses on the Cardiovascular System: A Review. JAMA Cardiology. 2020.

18. Heymann DL, Shindo N, Scientific WHO, Technical Advisory Group for Infectious H. COVID-19: what is next for public health? Lancet. 2020;395(10224):542–5.

19. Wan X, Wang W, Liu J, Tong T. Estimating the sample mean and standard deviation from the sample size, median, range and/or interquartile range. BMC Medical Research Methodology. 2014;14(1):135.

20. Stang A. Critical evaluation of the Newcastle-Ottawa scale for the assessment of the quality of nonrandomized studies in meta-analyses. European journal of epidemiology. 2010;25(9):603–5.

21. Argenziano MG, Bruce SL, Slater CL, Tiao JR, Baldwin MR, Barr RG, et al. Characterization and clinical course of 1000 Patients with COVID-19 in New York: retrospective case series. medRxiv. 2020:2020.04.20.20072116.

22. Bonetti G, Manelli F, Patroni A, Bettinardi A, Borrelli G, Fiordalisi G, et al. Laboratory predictors of death from coronavirus disease 2019 (COVID-19) in the area of Valcamonica, Italy. Clinical Chemistry and Laboratory Medicine. 2020.

23. Cao J, Hu X, Cheng W, Yu L, Tu WJ, Liu Q. Clinical features and short-term outcomes of 18 patients with corona virus disease 2019 in intensive care unit. Intensive Care Medicine. 2020;46(5):851–3.

24. Cao M, Zhang D, Wang Y, Lu Y, Zhu X, Li Y, et al. Clinical Features of Patients Infected with the 2019 Novel Coronavirus (COVID-19) in Shanghai, China. medRxiv. 2020.

25. Chen C, Yan J, Zhou N, Zhao J, Wang D. Analysis of myocardial injury in patients with COVID-19 and association between concomitant cardiovascular diseases and severity of COVID-19. Zhonghua xin xue guan bing za zhi. 2020;48:E008–E.

26. Chen C, Yi ZJ, Chang L, Shuo HZ, Ming Z, Pei T, et al. The characteristics and death risk factors of 132 COVID-19 pneumonia patients with comorbidities: a retrospective single center analysis in Wuhan, China. medRxiv. 2020:2020.05.07.20092882.

27. Chen R, Liang W, Jiang M, Guan W, Zhan C, Wang T, et al. Risk factors of fatal outcome in hospitalized subjects with coronavirus disease 2019 from a nationwide analysis in China. Chest. 2020.

28. Chen T, Wu D, Chen H, Yan W, Yang D, Chen G, et al. Clinical characteristics of 113 deceased patients with coronavirus disease 2019: retrospective study. BMJ. 2020;368.

29. Chen X, Zheng F, Qing Y, Ding S, Yang D, Lei C, et al. Epidemiological and clinical features of 291 cases with coronavirus disease 2019 in areas adjacent to Hubei, China: a double-center observational study. medRxiv. 2020.

30. Conversano A, Melillo F, Napolano A, Fominskiy E, Spessot M, Ciceri F, et al. RAAs inhibitors and outcome in patients with SARS-CoV-2 pneumonia. A case series study. Hypertension. 2020.

31. Deng Q, Hu B, Zhang Y, Wang H, Zhou X, Hu W, et al. Suspected myocardial injury in patients with COVID-19: Evidence from front-line clinical observation in Wuhan, China. Int J Cardiol. 2020.

32. Deng Y, Liu W, Liu K, Fang YY, Shang J, Zhou L, et al. Clinical characteristics of fatal and recovered cases of coronavirus disease 2019 (COVID-19) in Wuhan, China: a retrospective study. Chinese medical journal. 2020.

33. Du RH, Liang LR, Yang CQ, Wang W, Cao TZ, Li M, et al. Predictors of mortality for patients with COVID-19 pneumonia caused by SARS-CoV-2: a prospective cohort study. Eur Respir J. 2020;55(5).

34. Du RH, Liu LM, Yin W, Wang W, Guan LL, Yuan ML, et al. Hospitalization and Critical Care of 109 Decedents with COVID-19 Pneumonia in Wuhan, China Ann Am Thorac Soc. 2020.

35. Ebinger JE, Achamallah N, Ji H, Claggett BL, Sun N, Botting P, et al. Pre-Existing Traits Associated with Covid-19 Illness Severity. medRxiv. 2020:2020.04.29.20084533.

36. Fan H, Zhang L, Huang B, Zhu M, Zhou Y, Zhang H, et al. Cardiac injuries in patients with coronavirus disease 2019: Not to be ignored. International Journal of Infectious Diseases. 2020;96:294–7.

37. Ferguson J, Rosser JI, Quintero O, Scott J, Subramanian A, Gumma M, et al. Characteristics and Outcomes of Coronavirus Disease Patients under Nonsurge Conditions, Northern California, USA, March-April 2020. Emerg Infect Dis. 2020;26(8).

38. Fu L, Fei J, Xiang H-X, Xiang Y, Tan Z-X, Li M-D, et al. Influence factors of death risk among COVID-19 patients in Wuhan, China: a hospital-based case-cohort study. medRxiv. 2020.

39. Gagiannis D, Steinestel J, Hackenbroch C, Hannemann M, Umathum VG, Gebauer N, et al. COVID-19-induced acute respiratory failure: an exacerbation of organ-specific autoimmunity? medRxiv. 2020:2020.04.27.20077180.

40. Gaibazzi N, Martini C, Mattioli M, Tuttolomondo D, Guidorossi A, Suma S, et al. Lung disease severity, Coronary Artery Calcium, Coronary inflammation and Mortality in Coronavirus Disease 2019. medRxiv. 2020:2020.05.01.20087114.

41. Ge H, Zhu M, Du J, Zhou Y, Wang W, Zhang W, et al. Cardiac Structural and Functional Characteristics in Patients with Coronavirus Disease 2019: A Serial Echocardiographic Study. medRxiv. 2020:2020.05.12.20095885.

42. Gu T, Chu Q, Yu Z, Fa B, Li A, Xu L, et al. History of coronary heart disease increases the mortality rate of COVID-19 patients: a nested case-control study. medRxiv. 2020:2020.03.23.20041848.

43. Guan WJ, Liang WH, Zhao Y, Liang HR, Chen ZS, Li YM, et al. Comorbidity and its impact on 1590 patients with COVID-19 in China: a nationwide analysis. Eur Respir J. 2020;55(5).

44. He X, Lai J, Cheng J, Wang M, Liu Y, Xiao Z, et al. Impact of complicated myocardial injury on the clinical outcome of severe or critically ill COVID-19 patients. Zhonghua xin xue Guan Bing za zhi. 2020;48:E011–E.

45. Hong KS, Lee KH, Chung JH, Shin KC, Choi EY, Jin HJ, et al. Clinical Features and Outcomes of 98 Patients Hospitalized with SARS-CoV-2 Infection in Daegu, South Korea: A Brief Descriptive Study. Yonsei Med J. 2020;61(5):431–7.

46. Huang C, Wang Y, Li X, Ren L, Zhao J, Hu Y, et al. Clinical features of patients infected with 2019 novel coronavirus in Wuhan, China. The Lancet. 2020;395(10223):497–506.

47. Ketcham SW, Adie SK, Malliett A, Abdul-Aziz AA, Bitar A, Grafton G, et al. Coronavirus Disease-2019 in Heart Transplant Recipients in Southeastern Michigan: A Case Series. J Card Fail. 2020.

48. Lagi F, Piccica M, Graziani L, Vellere I, Botta A, Tilli M, et al. Early experience of an infectious and tropical diseases unit during the coronavirus disease (COVID-19) pandemic, Florence, Italy, February to March 2020. Eurosurveillance. 2020;25(17).

49. Levy TJ, Richardson S, Coppa K, Barnaby DP, McGinn T, Becker LB, et al. Estimating Survival of Hospitalized COVID-19 Patients from Admission Information. medRxiv. 2020:2020.04.22.20075416.

50. Li K, Wu J, Wu F, Guo D, Chen L, Fang Z, et al. The clinical and chest CT features associated with severe and critical COVID-19 pneumonia. Investigative radiology. 2020.

51. Liu Q, Fang X, Tokuno S, Chung U, Chen X, Dai X, et al. Prediction of the clinical outcome of COVID-19 patients using T lymphocyte subsets with 340 cases from Wuhan, China: a retrospective cohort study and a web visualization tool. medRxiv. 2020:2020.04.06.20056127.

52. Lodigiani C, Iapichino G, Carenzo L, Cecconi M, Ferrazzi P, Sebastian T, et al. Venous and arterial thromboembolic complications in COVID-19 patients admitted to an academic hospital in Milan, Italy. Thrombosis Research. 2020;191:9–14.

53. Ni W, Yang X, Liu J, Bao J, Li R, Xu Y, et al. Acute Myocardial Injury at Hospital Admission is Associated with All-cause Mortality in COVID-19. Journal of the American College of Cardiology. 2020.

54. Nikpouraghdam M, Jalali Farahani A, Alishiri G, Heydari S, Ebrahimnia M, Samadinia H, et al. Epidemiological characteristics of coronavirus disease 2019 (COVID-19) patients in IRAN: A single center study. J Clin Virol. 2020;127:104378.

55. Peng Y, Meng K, Guan H, Leng L, Zhu R, Wang B, et al. Clinical characteristics and outcomes of 112 cardiovascular disease patients infected by 2019-nCoV. Zhonghua xin xue guan bing za zhi. 2020;48:E004.

56. Rentsch CT, Kidwai-Khan F, Tate JP, Park LS, King JT, Skanderson M, et al. Covid-19 Testing, Hospital Admission, and Intensive Care Among 2,026,227 United States Veterans Aged 54-75 Years. medRxiv. 2020:2020.04.09.20059964.

57. Sapey E, Gallier S, Mainey C, Nightingale P, McNulty D, Crothers H, et al. Ethnicity and risk of death in patients hospitalised for COVID-19 infection: an observational cohort study in an urban catchment area. medRxiv. 2020:2020.05.05.20092296.

58. Shekhar R, Upadhyay S, Sheikh A, Atencio J, Kapuria D. Early experience with COVD-19 patients at tertiary care teaching hospital in southwestern United states. medRxiv. 2020:2020.05.15.20094284.

59. Shi S, Qin M, Cai Y, Liu T, Shen B, Yang F, et al. Characteristics and clinical significance of myocardial injury in patients with severe coronavirus disease 2019. Eur Heart J. 2020.

60. Sun H, Ning R, Tao Y, Yu C, Deng X, Zhao C, et al. Risk Factors for Mortality in 244 Older Adults With COVID-19 in Wuhan, China: A Retrospective Study. Journal of the American Geriatrics Society. 2020.

61. Tai S, Tang J, Yu B, Tang L, Wang Y, Zhang H, et al. Association between Cardiovascular Burden and Requirement of Intensive Care among Patients with Mild COVID-19. medRxiv. 2020:2020.05.25.20111757.

62. Wang D, Hu B, Hu C, Zhu F, Liu X, Zhang J, et al. Clinical Characteristics of 138 Hospitalized Patients with 2019 Novel Coronavirus-Infected Pneumonia in Wuhan, China. JAMA - Journal of the American Medical Association. 2020;323(11):1061–9.

63. Wang D, Yin Y, Hu C, Liu X, Zhang X, Zhou S, et al. Clinical course and outcome of 107 patients infected with the novel coronavirus, SARS-CoV-2, discharged from two hospitals in Wuhan, China. Critical Care. 2020;24(1).

64. Wang K, Zuo P, Liu Y, Zhang M, Zhao X, Xie S, et al. Clinical and laboratory predictors of in-hospital mortality in patients with COVID-19: a cohort study in Wuhan, China. Clin Infect Dis. 2020.

65. Wang L, He W, Yu X, Hu D, Bao M, Liu H, et al. Coronavirus disease 2019 in elderly patients: Characteristics and prognostic factors based on 4-week follow-up. J Infect. 2020;80(6):639–45.

66. Wang L, He WB, Yu XM, Liu HF, Zhou WJ, Jiang H. [Prognostic value of myocardial injury in patients with COVID-19]. Zhonghua Yan Ke Za Zhi. 2020;56(0):E009.

67. Yang X, Yu Y, Xu J, Shu H, Liu H, Wu Y, et al. Clinical course and outcomes of critically ill patients with SARS-CoV-2 pneumonia in Wuhan, China: a single-centered, retrospective, observational study. The Lancet Respiratory Medicine. 2020.

68. Yao Q, Wang P, Wang X, Qie G, Meng M, Tong X, et al. Retrospective study of risk factors for severe SARS-Cov-2 infections in hospitalized adult patients. Pol Arch Intern Med. 2020.

69. Zhang F, Yang D, Li J, Gao P, Chen T, Cheng Z, et al. Myocardial injury is associated with in-hospital mortality of confirmed or suspected COVID-19 in Wuhan, China: A single center retrospective cohort study. medRxiv. 2020.

70. Zhang J, Ding D, Cao C, Zhang J, Huang X, Fu P, et al. Myocardial characteristics as the prognosis for COVID-19 patients. medRxiv. 2020:2020.05.06.20068882.

71. Zhang J, Lu S, Wang X, Jia X, Li J, Lei H, et al. Do underlying cardiovascular diseases have any impact on hospitalised patients with COVID-19? Heart. 2020.

72. Zhou F, Yu T, Du R, Fan G, Liu Y, Liu Z, et al. Clinical course and risk factors for mortality of adult inpatients with COVID-19 in Wuhan, China: a retrospective cohort study. The Lancet. 2020;395(10229):1054–62.

73. Zhou X, Zhu J, Xu T. Clinical characteristics of coronavirus disease 2019 (COVID-19) patients with hypertension on renin-angiotensin system inhibitors. Clin Exp Hypertens. 2020:1–5.

74. Zou X, Li S, Fang M, Hu M, Bian Y, Ling J, et al. Acute Physiology and Chronic Health Evaluation II Score as a Predictor of Hospital Mortality in Patients of Coronavirus Disease 2019. Critical care medicine. 2020.

75. Zheng Y, Xu H, Yang M, Zeng Y, Chen H, Liu R, et al. Epidemiological characteristics and clinical features of 32 critical and 67 noncritical cases of COVID-19 in Chengdu. J Clin Virol. 2020;127:104366.

76. Li J, Song H, Hu Z. Clinical features and outcomes of 2019 novel coronavirus-infected patients with cardiac injury. medRxiv. 2020.

77. Arentz M, Yim E, Klaff L, Lokhandwala S, Riedo FX, Chong M, et al. Characteristics and Outcomes of 21 Critically Ill Patients with COVID-19 in Washington State. JAMA - Journal of the American Medical Association. 2020;323(16):1612–4.

78. Barrasa H, Rello J, Tejada S, Martín A, Balziskueta G, Vinuesa C, et al. SARS-CoV-2 in Spanish Intensive Care Units: Early experience with 15-day survival in Vitoria. Anaesthesia Critical Care and Pain Medicine. 2020.

79. Benetti E, Giliberti A, Emiliozzi A, Velentino F, Bergantini L, Fallerini C, et al. Clinical and molecular characterization of COVID-19 hospitalized patients. medRxiv. 2020:2020.05.22.20108845.

80. Bhayana R, Som A, Li MD, Carey DE, Anderson MA, Blake MA, et al. Abdominal Imaging Findings in COVID-19: Preliminary Observations. Radiology. 2020:201908.

81. Bradley BT, Maioli H, Johnston R, Chaudhry I, Fink SL, Xu H, et al. Histopathology and Ultrastructural Findings of Fatal COVID-19 Infections. medRxiv. 2020:2020.04.17.20058545.

82. Brat GA, Weber GM, Gehlenborg N, Avillach P, Palmer NP, Chiovato L, et al. International Electronic Health Record-Derived COVID-19 Clinical Course Profiles: The 4CE Consortium. medRxiv. 2020:2020.04.13.20059691.

83. Bravi F, Flacco ME, Carradori T, Volta CA, Cosenza G, De Togni A, et al. Predictors of severe or lethal COVID-19, including Angiotensin Converting Enzyme Inhibitors and Angiotensin II Receptor Blockers, in a sample of infected Italian citizens. medRxiv. 2020:2020.05.21.20109082.

84. Buckner FS, McCulloch DJ, Atluri V, Blain M, McGuffin SA, Nalla AK, et al. Clinical Features and Outcomes of 105 Hospitalized patients with COVID-19 in Seattle, Washington. Clin Infect Dis. 2020.

85. Cai Q, Chen F, Wang T, Luo F, Liu X, Wu Q, et al. Obesity and COVID-19 Severity in a Designated Hospital in Shenzhen, China. Diabetes Care. 2020.

86. Cai Q, Huang D, Ou P, Yu H, Zhu Z, Xia Z, et al. COVID-19 in a designated infectious diseases hospital outside Hubei Province, China. Allergy: European Journal of Allergy and Clinical Immunology. 2020.

87. Casas Rojo JM, Antón Santos JM,Millán Núñez-Cortés J, Lumbreras Bermejo C, Ramos Rincón JM, Roy-Vallejo E, et al. Clinical characteristics of patients hospitalized with COVID-19 in Spain: results from the SEMI-COVID-19 Network. medRxiv. 2020:2020.05.24.20111971.

88. Chen G, Wu D, Guo W, Cao Y, Huang D, Wang H, et al. Clinical and immunological features of severe and moderate coronavirus disease 2019. Journal of Clinical Investigation. 2020;130(5):2620–9.

89. Chen J, Fan H, Zhang L, Huang B, Zhu M, Zhou Y, et al. Retrospective analysis of clinical features in 101 death cases with COVID-19. medRxiv. 2020.

90. Chen J, Xu X, Hu J, Chen Q, Xu F, Liang H, et al. Clinical Course and Risk Factors for Recurrence of Positive SARS-CoV-2 RNA: A Retrospective Cohort Study from Wuhan, China. medRxiv. 2020:2020.05.08.20095018.

91. Chen N, Zhou M, Dong X, Qu J, Gong F, Han Y, et al. Epidemiological and clinical characteristics of 99 cases of 2019 novel coronavirus pneumonia in Wuhan, China: a descriptive study. The Lancet. 2020;395(10223):507–13.

92. Chen N, Zhou M, Dong X, Qu J, Gong F, Han Y, et al. Epidemiological and clinical characteristics of 99 cases of 2019 novel coronavirus pneumonia in Wuhan, China: a descriptive study. The Lancet. 2020;395(10223):507–13.

93. Chen Q, Zheng Z, Zhang C, Zhang X, Wu H, Wang J, et al. Clinical characteristics of 145 patients with corona virus disease 2019 (COVID-19) in Taizhou, Zhejiang, China. Infection. 2020:1–9.

94. Cui S, Chen S, Li X, Liu S, Wang F. Prevalence of venous thromboembolism in patients with severe novel coronavirus pneumonia. Journal of Thrombosis and Haemostasis. 2020.

95. Cummings MJ, Baldwin MR, Abrams D, Jacobson SD, Meyer BJ, Balough EM, et al. Epidemiology, clinical course, and outcomes of critically ill adults with COVID-19 in New York City: a prospective cohort study. Lancet. 2020.

96. Dai H, Zhang X, Xia J, Zhang T, Shang Y, Huang R, et al. High-resolution Chest CT Features and Clinical Characteristics of Patients Infected with COVID-19 in Jiangsu, China. International Journal of Infectious Diseases. 2020;95:106–12.

97. Demelo-Rodríguez P, Cervilla-Muñoz E, Ordieres-Ortega L, Parra-Virto A, Toledano-Macías M, Toledo-Samaniego N, et al. Incidence of asymptomatic deep vein thrombosis in patients with COVID-19 pneumonia and elevated D-dimer levels. Thrombosis Research. 2020;192:23–6.

98. Du Y, Tu L, Zhu P, Mu M, Wang R, Yang P, et al. Clinical Features of 85 Fatal Cases of COVID-19 from Wuhan: A Retrospective Observational Study. Am J Respir Crit Care Med. 2020.

99. Estebanez M, Ramirez-Olivencia G, Mata T, Marti D, Gutierrez C, De Dios B, et al. Clinical evaluation of IFN beta1b in COVID-19 pneumonia: a retrospective study. medRxiv. 2020:2020.05.15.20084293.

100. Fei J, Fu L, Li Y, Xiang H-X, Xiang Y, Li M-D, et al. Reduction of lymphocyte at early stage elevates severity and death risk of COVID-19 patients: a hospital-based case-cohort study. medRxiv. 2020:2020.04.02.20050955.

101. Feng X, Li P, Ma L, Liang H, Lei J, Li W, et al. Clinical Characteristics and Short-Term Outcomes of Severe Patients with COVID-19 in Wuhan, China. medRxiv. 2020:2020.04.24.20078063.

102. Fogarty H, Townsend L, Ni Cheallaigh C, Bergin C, Martin-Loeches I, Browne P, et al. COVID-19 Coagulopathy in Caucasian patients. British Journal of Haematology. 2020.

103. Fu J, Kong J, Wang W, Wu M, Yao L, Wang Z, et al. The clinical implication of dynamic neutrophil to lymphocyte ratio and D-dimer in COVID-19: A retrospective study in Suzhou China. Thrombosis Research. 2020;192:3–8.

104. Gao L, Jiang D, Wen XS, Cheng XC, Sun M, He B, et al. Prognostic value of NT-proBNP in patients with severe COVID-19. Respiratory Research. 2020;21(1).

105. Garcia-Olivé I, Sintes H, Radua J, Abad Capa J, Rosell A. D-dimer in patients infected with COVID-19 and suspected pulmonary embolism. Respiratory Medicine. 2020;169.

106. Giorgi Rossi P, Marino M, Formisano D, Venturelli F, Vicentini M, Grilli R. Characteristics and outcomes of a cohort of SARS-CoV-2 patients in the Province of Reggio Emilia, Italy. medRxiv. 2020:2020.04.13.20063545.

107. Grasselli G, Zangrillo A, Zanella A, Antonelli M, Cabrini L, Castelli A, et al. Baseline Characteristics and Outcomes of 1591 Patients Infected With SARS-CoV-2 Admitted to ICUs of the Lombardy Region, Italy. Jama. 2020;323(16):1574–81.

108. Han H, Xie L, Liu R, Yang J, Liu F, Wu K, et al. Analysis of heart injury laboratory parameters in 273 COVID-19 patients in one hospital in Wuhan, China. Journal of Medical Virology. 2020.

109. Han Y, Zhang H, Mu S, Wei W, Jin C, Xue Y, et al. Lactate dehydrogenase, a Risk Factor of Severe COVID-19 Patients. medRxiv. 2020:2020.03.24.20040162.

110. He R, Lu Z, Zhang L, Fan T, Xiong R, Shen X, et al. The clinical course and its correlated immune status in COVID-19 pneumonia. Journal of Clinical Virology. 2020;127.

111. Helms J, Tacquard C, Severac F, Leonard-Lorant I, Ohana M, Delabranche X, et al. High risk of thrombosis in patients with severe SARS-CoV-2 infection: a multicenter prospective cohort study. Intensive Care Medicine. 2020.

112. Hong N, Yu W, Xia J, Shen Y, Yap M, Han W. Evaluation of ocular symptoms and tropism of SARS-CoV-2 in patients confirmed with COVID-19. Acta Ophthalmol. 2020.

113. Huang J, Cheng A, Lin S, Zhu Y, Chen G. Individualized prediction nomograms for disease progression in mild COVID-19. Journal of Medical Virology.2020.

114. Hui H, Zhang Y, Yang X, Wang X, He B, Li L, et al. Clinical and radiographic features of cardiac injury in patients with 2019 novel coronavirus pneumonia. MedRxiv. 2020.

115. Inciardi RM, Adamo M, Lupi L, Cani DS, Di Pasquale M, Tomasoni D, et al. Characteristics and outcomes of patients hospitalized for COVID-19 and cardiac disease in Northern Italy. Eur Heart J. 2020;41(19):1821–9.

116. Itelman E, Wasserstrum Y, Segev A, Avaky C, Negru L, Cohen D, et al. Clinical Characterization of 162 COVID-19 patients in Israel: Preliminary Report from a Large Tertiary Center. Isr Med Assoc J. 2020;22(5):271–4.

117. Jain S, Workman V, Ganeshan R, Obasare ER, Burr A, DeBiasi RM, et al. ENHANCED ECG MONITORING OF COVID-19 PATIENTS. Heart rhythm. 2020.

118. Jeong EK, Park O, Park YJ, Park SY, Kim YM, Kim J, et al. Coronavirus disease-19: The first 7,755 cases in the Republic of Korea. Osong Public Health and Research Perspectives. 2020;11(2):85–90.

119. Khera R, Clark C, Lu Y, Guo Y, Ren S, Truax B, et al. Association of Angiotensin-Converting Enzyme Inhibitors and Angiotensin Receptor Blockers with the Risk of Hospitalization and Death in Hypertensive Patients with Coronavirus Disease-19. medRxiv. 2020:2020.05.17.20104943.

120. Kim L, Garg S, O’Halloran A, Whitaker M, Pham H, Anderson EJ, et al. Interim Analysis of Risk Factors for Severe Outcomes among a Cohort of Hospitalized Adults Identified through the U.S. Coronavirus Disease 2019 (COVID-19)-Associated Hospitalization Surveillance Network (COVID-NET). medRxiv. 2020:2020.05.18.20103390.

121. Klok FA, Kruip Mjha, van der Meer Njm, Arbous MS, Gommers Dampj, Kant KM, et al. Incidence of thrombotic complications in critically ill ICU patients with COVID-19. Thrombosis Research. 2020.

122. Kolin DA, Kulm S, Elemento O. Clinical and Genetic Characteristics of Covid-19 Patients from UK Biobank. medRxiv. 2020:2020.05.05.20075507.

123. Kuno T, Takahashi M, Obata R, Maeda T. Cardiovascular comorbidities, cardiac injury and prognosis of COVID-19 in New York City. American heart journal. 2020.

124. Lala A, Johnson KW, Russak AJ, Paranjpe I, Zhao S, Solani S, et al. Prevalence and Impact of Myocardial Injury in Patients Hospitalized with COVID-19 Infection. medRxiv. 2020:2020.04.20.20072702.

125. Latif F, Farr MA, Clerkin KJ, Habal MV, Takeda K, Naka Y, et al. Characteristics and Outcomes of Recipients of Heart Transplant With Coronavirus Disease 2019. JAMA Cardiol. 2020.

126. Lax SF, Skok K, Zechner P, Kessler HH, Kaufmann N, Koelblinger C, et al. Pulmonary Arterial Thrombosis in COVID-19 With Fatal Outcome: Results From a Prospective, Single-Center, Clinicopathologic Case Series. Annals of internal medicine. 2020.

127. Li M, Dong Y, Wang H, Guo W, Zhou H, Zhang Z, et al. Cardiovascular disease potentially contributes to the progression and poor prognosis of COVID-19. Nutr Metab Cardiovasc Dis. 2020.

128. Li R, Tian J, Yang F, Lv L, Yu J, Sun G, et al. Clinical characteristics of 225 patients with COVID-19 in a tertiary Hospital near Wuhan, China. J Clin Virol. 2020;127:104363.

129. Li X, Xu S, Yu M, Wang K, Tao Y, Zhou Y, et al. Risk factors for severity and mortality in adult COVID-19 inpatients in Wuhan. J Allergy Clin Immunol. 2020.

130. Lian J, Jin X, Hao S, Cai H, Zhang S, Zheng L, et al. Analysis of Epidemiological and Clinical features in older patients with Corona Virus Disease 2019 (COVID-19) out of Wuhan. Clinical infectious diseases: an official publication of the Infectious Diseases Society of America. 2020.

131. Lian J, Jin X, Hao S, Jia H, Cai H, Zhang X, et al. Epidemiological, clinical, and virological characteristics of 465 hospitalized cases of coronavirus disease 2019 (COVID-19) from Zhejiang province in China. Influenza Other Respir Viruses. 2020.

132. Liu F, Li L, Xu M, Wu J, Luo D, Zhu Y, et al. Prognostic value of interleukin-6, C-reactive protein, and procalcitonin in patients with COVID-19. Journal of Clinical Virology. 2020;127.

133. Liu F, Xu A, Zhang Y, Xuan W, Yan T, Pan K, et al. Patients of COVID-19 may benefit from sustained lopinavir-combined regimen and the increase of eosinophil may predict the outcome of COVID-19 progression. International Journal of Infectious Diseases. 2020.

134. Liu K, Fang Y-Y, Deng Y, Liu W, Wang M-F, Ma J-P, et al. Clinical characteristics of novel coronavirus cases in tertiary hospitals in Hubei Province. Chinese medical journal. 2020;133(9):1025–31.

135. Liu R, Ming X, Zhu H, Song L, Gao Z, Gao L, et al. Association of cardiovascular manifestations with in-hospital outcomes in patients with COVID-19: a hospital staff data. MedRxiv. 2020.

136. Liu Y, Du X, Chen J, Jin Y, Peng L, Wang HHX, et al. Neutrophil-to-lymphocyte ratio as an independent risk factor for mortality in hospitalized patients with COVID-19. Journal of Infection. 2020.

137. Lu L, Xiong W, Liu D, Liu J, Yang D, Li N, et al. New-onset acute symptomatic seizure and risk factors in Corona Virus Disease 2019: A Retrospective Multicenter Study. Epilepsia. 2020.

138. Mao L, Jin H, Wang M, Hu Y, Chen S, He Q, et al. Neurologic Manifestations of Hospitalized Patients With Coronavirus Disease 2019 in Wuhan, China. JAMA Neurol. 2020.

139. Menter T, Haslbauer JD, Nienhold R, Savic S, Hopfer H, Deigendesch N, et al. Post-mortem examination of COVID19 patients reveals diffuse alveolar damage with severe capillary congestion and variegated findings of lungs and other organs suggesting vascular dysfunction. Histopathology. 2020.

140. Miao C, Zhuang J, Jin M, Xiong H, Huang P, Zhao Q, et al. A comparative multi-centre study on the clinical and imaging features of comfirmed and uncomfirmed patients with COVID-19. medRxiv. 2020.

141. Middeldorp S, Coppens M, van Haaps TF, Foppen M, Vlaar AP, Müller MCA, et al. Incidence of venous thromboembolism in hospitalized patients with COVID-19. Journal of thrombosis and haemostasis: JTH. 2020.

142. Mo P, Xing Y, Xiao Y, Deng L, Zhao Q, Wang H, et al. Clinical characteristics of refractory COVID-19 pneumonia in Wuhan, China. Clinical Infectious Diseases. 2020.

143. Murk JL, van de Biggelaar R, Stohr J, Verweij J, Buiting A, Wittens S, et al. [The first 100 COVID-19 patients admitted to the Elisabeth-Tweesteden Hospital, Tilburg, The Netherlands]. Ned Tijdschr Geneeskd. 2020;164.

144. Nie S, Zhao X, Zhao K, Zhang Z, Zhang Z, Zhang Z. Metabolic disturbances and inflammatory dysfunction predict severity of coronavirus disease 2019 (COVID-19): a retrospective study. medRxiv. 2020.

145. Niu S, Tian S, Lou J, Kang X, Zhang L, Lian H, et al. Clinical characteristics of older patients infected with COVID-19: A descriptive study. Archives of Gerontology and Geriatrics. 2020;89.

146. Nunes Duarte-Neto A, de Almeida Monteiro RA, da Silva LFF, Malheiros D, de Oliveira EP, Theodoro Filho J, et al. Pulmonary and systemic involvement of COVID-19 assessed by ultrasound-guided minimally invasive autopsy. Histopathology. 2020.

147. Öztürk F, Karaduman M, Çoldur R, İncecik Ş, Güneş Y, Tuncer M. Interpretation of arrhythmogenic effects of COVID-19 disease through ECG. Aging Male. 2020:1–4.

148. Palaiodimos L, Kokkinidis DG, Li W, Karamanis D, Ognibene J, Arora S, et al. Severe obesity is associated with higher in-hospital mortality in a cohort of patients with COVID-19 in the Bronx, New York. Metabolism: Clinical and Experimental. 2020;108.

149. Pavoni V, Gianesello L, Pazzi M, Stera C, Meconi T, Frigieri FC. Evaluation of coagulation function by rotation thromboelastometry in critically ill patients with severe COVID-19 pneumonia. Journal of Thrombosis and Thrombolysis. 2020.

150. Petrilli CM, Jones SA, Yang J, Rajagopalan H, O’Donnell L, Chernyak Y, et al. Factors associated with hospital admission and critical illness among 5279 people with coronavirus disease 2019 in New York City: prospective cohort study. Bmj. 2020;369:m1966.

151. Piva S, Filippini M, Turla F, Cattaneo S, Margola A, De Fulviis S, et al. Clinical presentation and initial management critically ill patients with severe acute respiratory syndrome coronavirus 2 (SARS-CoV-2) infection in Brescia, Italy. J Crit Care. 2020;58:29–33.

152. Poyiadji N, Cormier P, Patel PY, Hadied MO, Bhargava P, Khanna K, et al. Acute Pulmonary Embolism and COVID-19. Radiology. 2020:201955.

153. Qi D, Yan X, Tang X, Peng J, Yu Q, Feng L, et al. Epidemiological and clinical features of 2019-nCoV acute respiratory disease cases in Chongqing municipality, China: a retrospective, descriptive, multiple-center study. medRxiv. 2020.

154. Qian G-Q, Yang N-B, Ding F, Ma AHY, Wang Z-Y, Shen Y-F, et al. Epidemiologic and Clinical Characteristics of 91 Hospitalized Patients with COVID-19 in Zhejiang, China: A retrospective, multi-centre case series. QJM: An International Journal of Medicine. 2020.

155. Razavi AC, Kelly TN, He J, Fernandez C, Whelton PK, Krousel-Wood M, et al. Cardiovascular Disease Prevention and Implications of COVID-19: An Evolving Case Study in the Crescent City. J Am Heart Assoc. 2020:e016997.

156. Richardson S, Hirsch JS, Narasimhan M, Crawford JM, McGinn T, Davidson KW, et al. Presenting Characteristics, Comorbidities, and Outcomes among 5700 Patients Hospitalized with COVID-19 in the New York City Area. JAMA - Journal of the American Medical Association. 2020:E1–E8.

157. Ruan Q, Yang K, Wang W, Jiang L, Song J. Clinical predictors of mortality due to COVID-19 based on an analysis of data of 150 patients from Wuhan, China. Intensive Care Medicine. 2020;46(5):846–8.

158. Sami R, Soltaninejad F, Amra B, Naderi Z, Haghjooy Javanmard S, Iraj B, et al. A one-year hospital-based prospective COVID-19 open-cohort in the Eastern Mediterranean region: The Khorshid COVID Cohort (KCC) study. medRxiv. 2020:2020.05.11.20096727.

159. Secco GG, Tarantini G, Mazzarotto P, Garbo R, Parisi R, Maggio S, et al. Invasive strategy for COVID patients presenting with acute coronary syndrome: The first multicenter Italian experience. Catheter Cardiovasc Interv. 2020.

160. Shao F, Xu S, Ma X, Xu Z, Lyu J, Ng M, et al. In-hospital cardiac arrest outcomes among patients with COVID-19 pneumonia in Wuhan, China. Resuscitation. 2020;151:18–23.

161. Shi H, Han X, Jiang N, Cao Y, Alwalid O, Gu J, et al. Radiological findings from 81 patients with COVID-19 pneumonia in Wuhan, China: a descriptive study. The Lancet Infectious Diseases. 2020.

162. Shi Q, Zhao K, Yu J, Feng J, Zhao K, Zhang X, et al. Clinical characteristics of 101 non-surviving hospitalized patients with COVID-19: A single center, retrospective study. medRxiv. 2020.

163. Shi S, Qin M, Shen B, Cai Y, Liu T, Yang F, et al. Association of Cardiac Injury with Mortality in Hospitalized Patients with COVID-19 in Wuhan, China. JAMA Cardiology. 2020.

164. Shi Y, Yu X, Zhao H, Wang H, Zhao R, Sheng J. Host susceptibility to severe COVID-19 and establishment of a host risk score: Findings of 487 cases outside Wuhan. Critical Care. 2020;24(1).

165. Simonnet A, Chetboun M, Poissy J, Raverdy V, Noulette J, Duhamel A, et al. High prevalence of obesity in severe acute respiratory syndrome coronavirus-2 (SARS-CoV-2) requiring invasive mechanical ventilation. Obesity. 2020.

166. Song F, Shi N, Shan F, Zhang Z, Shen J, Lu H, et al. Emerging 2019 novel coronavirus (2019-nCoV) pneumonia. Radiology. 2020;295(1):210–7.

167. Sun C, Zhang X, Dai Y, Xu X, Zhao J. Clinical analysis of 150 cases of 2019 novel coronavirus infection in Nanyang City, Henan Province. Zhonghua jie he he hu xi za zhi= Zhonghua Jiehe he Huxi Zazhi= Chinese Journal of Tuberculosis and Respiratory Diseases. 2020;43:E042–E.

168. Sun L, Shen L, Fan J, Gu F, Hu M, An Y, et al. Clinical Features of Patients with Coronavirus Disease 2019 (COVID-19) from a Designated Hospital in Beijing, China. J Med Virol. 2020.

169. Vahidy FS, Nicolas JC, Meeks JR, Khan O, Jones SL, Masud F, et al. Racial and Ethnic Disparities in SARS-CoV-2 Pandemic: Analysis of a COVID-19 Observational Registry for a Diverse U.S. Metropolitan Population. medRxiv. 2020:2020.04.24.20073148.

170. Vicenzi M, Di Cosola R, Ruscica M, Ratti A, Rota I, Rota F, et al. The liaison between respiratory failure and high blood pressure: evidence from COVID-19 patients. Eur Respir J. 2020.

171. Wan S, Xiang Y, Fang W, Zheng Y, Li B, Hu Y, et al. Clinical features and treatment of COVID-19 patients in northeast Chongqing. Journal of Medical Virology. 2020.

172. Wang K, Kang S, Tian R, Zhang X, Wang Y. Imaging manifestations and diagnostic value of chest CT of coronavirus disease 2019 (COVID-19) in the Xiaogan area. Clinical Radiology. 2020;75(5):341–7.

173. Wang W, He J, Wu S. The definition and risks of cytokine release syndrome-like in 11 COVID-19-infected pneumonia critically ill patients: disease characteristics and retrospective analysis. Medrxiv. 2020.

174. Wang X, Fang J, Zhu Y, Chen L, Ding F, Zhou R, et al. Clinical characteristics of non-critically ill patients with novel coronavirus infection (COVID-19) in a Fangcang Hospital. Clin Microbiol Infect. 2020.

175. Wang Z, Yang B, Li Q, Wen L, Zhang R. Clinical features of 69 cases with coronavirus disease 2019 in Wuhan, China. Clinical infectious diseases. 2020.

176. Wei JF, Huang FY, Xiong TY, Liu Q, Chen H, Wang H, et al. Acute myocardial injury is common in patients with covid-19 and impairs their prognosis. Heart. 2020.

177. Wright FL, Vogler TO, Moore EE, Moore HB, Wohlauer MV, Urban S, et al. Fibrinolysis shutdown correlates to thromboembolic events in severe COVID-19 Infection. Journal of the American College of Surgeons. 2020.

178. Wu C, Chen X, Cai Y, Xia J, Zhou X, Xu S, et al. Risk Factors Associated with Acute Respiratory Distress Syndrome and Death in Patients with Coronavirus Disease 2019 Pneumonia in Wuhan, China. JAMA Internal Medicine. 2020.

179. Wu C, Hu X, Song J, Du C, Xu J, Yang D, et al. Heart injury signs are associated with higher and earlier mortality in coronavirus disease 2019 (COVID-19). MedRxiv. 2020.

180. Wu J, Liu J, Zhao X, Liu C, Wang W, Wang D, et al. Clinical characteristics of imported cases of COVID-19 in Jiangsu province: a multicenter descriptive study. Clin Infect Dis. 2020;10.

181. Xi A, Zhuo M, Dai J, Ding Y, Ma X, Ma X, et al. Epidemiological and clinical characteristics of discharged patients infected with SARS-CoV-2 on the Qinghai plateau. J Med Virol. 2020.

182. Xu H, Hou K, Xu H, Li Z, Chen H, Zhang N, et al. Acute Myocardial Injury of Patients with Coronavirus Disease 2019. MedRxiv. 2020.

183. Xu K, Chen Y, Yuan J, Yi P, Ding C, Wu W, et al. Factors associated with prolonged viral RNA shedding in patients with COVID-19. Clin Infect Dis. 2020.

184. Xu S, Fu L, Fei J, Xiang H-X, Xiang Y, Tan Z-X, et al. Acute kidney injury at early stage as a negative prognostic indicator of patients with COVID-19: a hospital-based retrospective analysis. medRxiv. 2020.

185. Xu T, Chen C, Zhu Z, Cui M, Chen C, Dai H, et al. Clinical features and dynamics of viral load in imported and non-imported patients with COVID-19. International Journal of Infectious Diseases. 2020.

186. Xu X, Yu C, Qu J, Zhang L, Jiang S, Huang D, et al. Imaging and clinical features of patients with 2019 novel coronavirus SARS-CoV-2. European journal of nuclear medicine and molecular imaging. 2020:1–6.

187. Xu X-W, Wu X-X, Jiang X-G, Xu K-J, Ying L-J, Ma C-L, et al. Clinical findings in a group of patients infected with the 2019 novel coronavirus (SARS-Cov-2) outside of Wuhan, China: retrospective case series. bmj. 2020;368.

188. Yang AP, Liu JP, Tao WQ, Li HM. The diagnostic and predictive role of NLR, d-NLR and PLR in COVID-19 patients. International Immunopharmacology. 2020;84.

189. Yang F, Shi S, Zhu J, Shi J, Dai K, Chen X. Analysis of 92 deceased patients with COVID-19. Journal of Medical Virology. 2020.

190. Yang Z, Shi J, He Z, Lu Y, Xu Q, Ye C, et al. Predictors for imaging progression on chest ct from coronavirus disease 2019 (covid-19) patients. Aging. 2020;12(7):6037–48.

191. Yin L, Mou H, Shao J, Zhu Y, Pang X, Yang J, et al. Correlation between Heart fatty acid binding protein and severe COVID-19: A case-control study. PLoS ONE. 2020;15(4).

192. Yu C, Qu J, Zhang L, Jiang S, Chen B, Guan W, et al. High resolution CT findings and clinical features of COVID-19 in Guangzhou. Chinese Journal of Radiology (China). 2020;54(4):314–7.

193. Yu X, Sun X, Cui P, Pan H, Lin S, Han R, et al. Epidemiological and Clinical Characteristics of 333 Confirmed Cases with Coronavirus Disease 2019 in Shanghai, China. Transboundary and Emerging Diseases. 2020.

194. Yuan J, Zou R, Zeng L, Kou S, Lan J, Li X, et al. The correlation between viral clearance and biochemical outcomes of 94 COVID-19 infected discharged patients. Inflammation Research. 2020;69(6):599–606.

195. Zangrillo A, Beretta L, Scandroglio AM, Monti G, Fominskiy E, Colombo S, et al. Characteristics, treatment, outcomes and cause of death of invasively ventilated patients with COVID-19 ARDS in Milan, Italy. Crit Care Resusc. 2020.

196. Zeng L, Li J, Liao M, Hua R, Huang P, Zhang M, et al. Risk assessment of progression to severe conditions for patients with COVID-19 pneumonia: a single-center retrospective study. medRxiv. 2020.

197. Zhang B, Zhou X, Qiu Y, Feng F, Feng J, Jia Y, et al. Clinical characteristics of 82 death cases with COVID-19. MedRxiv. 2020.

198. Zhang G, Hu C, Luo L, Fang F, Chen Y, Li J, et al. Clinical features and short-term outcomes of 221 patients with COVID-19 in Wuhan, China. Journal of Clinical Virology. 2020;127.

199. Zhang J, Wang X, Jia X, Li J, Hu K, Chen G, et al. Risk factors for disease severity, unimprovement, and mortality in COVID-19 patients in Wuhan, China. Clinical Microbiology and Infection. 2020.

200. Zhang J, Yu M, Tong S, Liu LY, Tang LV. Predictive factors for disease progression in hospitalized patients with coronavirus disease 2019 in Wuhan, China. Journal of Clinical Virology. 2020;127.

201. Zhang J-j, Dong X, Cao Y-y, Yuan Y-d, Yang Y-b, Yan Y-q, et al. Clinical characteristics of 140 patients infected with SARS-CoV-2 in Wuhan, China. Allergy. 2020.

202. Zhang L, Feng X, Zhang D, Jiang C, Mei H, Wang J, et al. Deep Vein Thrombosis in Hospitalized Patients with Coronavirus Disease 2019 (COVID-19) in Wuhan, China: Prevalence, Risk Factors, and Outcome. Circulation. 2020.

203. Zhang X, Cai H, Hu J, Lian J, Gu J, Zhang S, et al. Epidemiological, clinical characteristics of cases of SARS-CoV-2 infection with abnormal imaging findings. International Journal of Infectious Diseases. 2020;94:81–7.

204. Zhang Y, Cui Y, Shen M, Zhang J, Liu B, Dai M, et al. Association of diabetes mellitus with disease severity and prognosis in COVID-19: A retrospective cohort study. Diabetes Research and Clinical Practice. 2020;165:108227.

205. Zhang Y, Deng A, Hu T, Chen X, Zhuang Y, Tan X, et al. Clinical outcomes of COVID-19 cases and influencing factors in Guangdong province. Zhonghua liu xing bing xue za zhi= Zhonghua liuxingbingxue zazhi. 2020;41:E057–E.

206. Zhao S, Ling K, Yan H, Zhong L, Peng X, Yao S, et al. Anesthetic management of patients with suspected 2019 novel coronavirus infection during emergency procedures. Journal of cardiothoracic and vascular anesthesia. 2020.

207. Zhao W, Zhong Z, Xie X, Yu Q, Liu J. Relation between chest CT findings and clinical conditions of coronavirus disease (covid-19) pneumonia: A multicenter study. American Journal of Roentgenology. 2020;214(5):1072–7.

208. Zhao XY, Xu XX, Yin HS, Hu QM, Xiong T, Tang YY, et al. Clinical characteristics of patients with 2019 coronavirus disease in a non-Wuhan area of Hubei Province, China: A retrospective study. BMC Infectious Diseases. 2020;20(1).

209. Zhao Z, Xie J, Yin M, Yang Y, He H, Jin T, et al. Clinical and laboratory profiles of 75 hospitalized patients with novel coronavirus disease 2019 in Hefei, China. MedRxiv. 2020.

210. Zheng F, Tang W, Li H, Huang YX, Xie YL, Zhou ZG. Clinical characteristics of 161 cases of corona virus disease 2019 (COVID-19) in Changsha. European Review for Medical and Pharmacological Sciences. 2020;24(6):3404–10.

211. Zheng Y, Sun LJ, Xu M, Pan J, Zhang YT, Fang XL, et al. Clinical characteristics of 34 COVID-19 patients admitted to intensive care unit in Hangzhou, China. J Zhejiang Univ Sci B. 2020;21(5):378–87.

212. Zhou B, She J, Wang Y, Ma X. The clinical characteristics of myocardial injury in severe and very severe patients with 2019 novel coronavirus disease. Journal of Infection. 2020.

213. Zhou F, Yu X, Tong X, Zhang R. Clinical features and outcomes of 197 adult discharged patients with COIVD-19 in Yichang, Hubei. medRxiv. 2020.

214. Zhou Y, He Y, Yang H, Yu H, Wang T, Chen Z, et al. Development and validation a nomogram for predicting the risk of severe COVID-19: A multi-center study in Sichuan, China. PLoS One. 2020;15(5):e0233328.

215. Zhou Z, Zhao N, Shu Y, Han S, Chen B, Shu X. Effect of Gastrointestinal Symptoms in Patients With COVID-19. Gastroenterology. 2020;158(8):2294–7.

216. Zhu Z, Cai T, Fan L, Lou K, Hua X, Huang Z, et al. Clinical value of immune-inflammatory parameters to assess the severity of coronavirus disease 2019. Int J Infect Dis. 2020;95:332–9.

217. Zhu Z, Tang J, Chai X, Fang Z, Liu Q, Hu X, et al. How to differentiate COVID-19 pneumonia from heart failure with computed tomography at initial medical contact during epidemic period. medRxiv. 2020.

218. Badawi A, Ryoo SG. Prevalence of comorbidities in the Middle East respiratory syndrome coronavirus (MERS-CoV): a systematic review and meta-analysis. International Journal of Infectious Diseases. 2016;49:129–33.

219. Xiong T-Y, Redwood S, Prendergast B, Chen M. Coronaviruses and the cardiovascular system: acute and long-term implications. Eur Heart J. 2020.

220. Zheng Y-Y, Ma Y-T, Zhang J-Y, Xie X. COVID-19 and the cardiovascular system. Nature Reviews Cardiology. 2020.

221. Bansal M. Cardiovascular disease and COVID-19. Diabetes & metabolic syndrome. 2020;14(3):247–50.

222. Clerkin KJ, Fried JA, Raikhelkar J, Sayer G, Griffin JM, Masoumi A, et al. COVID-19 and Cardiovascular Disease. Circulation. 2020;141(20):1648–55.

223. Bavishi C, Bonow RO, Trivedi V, Abbott JD, Messerli FH, Bhatt DL. Acute myocardial injury in patients hospitalized with COVID-19 infection: A review. Progress in cardiovascular diseases. 2020:S0033-620(20)30123-7.

224. Hoffmann M, Kleine-Weber H, Schroeder S, Krüger N, Herrler T, Erichsen S, et al. SARS-CoV-2 Cell Entry Depends on ACE2 and TMPRSS2 and Is Blocked by a Clinically Proven Protease Inhibitor. Cell. 2020.

225. Zhu H, Rhee J-W, Cheng P, Waliany S, Chang A, Witteles RM, et al. Cardiovascular Complications in Patients with COVID-19: Consequences of Viral Toxicities and Host Immune Response. Current cardiology reports. 2020;22(5):32-.

226. Connors JM, Levy JH. COVID-19 and its implications for thrombosis and anticoagulation. Blood. 2020;135(23):2033–40.

227. Clerkin KJ, Fried JA, Raikhelkar J, Sayer G, Griffin JM, Masoumi A, et al. Coronavirus Disease 2019 (COVID-19) and Cardiovascular Disease. Circulation. 2020.

228. South AM, Diz D, Chappell MC. COVID-19, ACE2 and the Cardiovascular Consequences. Am J Physiol Heart Circ Physiol. 2020.

229. Zheng Y-Y, Ma Y-T, Zhang J-Y, Xie X.COVID-19 and the cardiovascular system. Nature Reviews Cardiology. 2020;17(5):259–60.

230. Schiffrin EL, Flack JM, Ito S, Muntner P, Webb RC. Hypertension and COVID-19. American journal of hypertension. 2020;33(5):373–4.

231. Fang L, Karakiulakis G, Roth M. Are patients with hypertension and diabetes mellitus at increased risk for COVID-19 infection? The Lancet Respiratory medicine. 2020;8(4):e21–e.

232. AlGhatrif M, Cingolani O, Lakatta EG. The Dilemma of Coronavirus Disease 2019, Aging, and Cardiovascular Disease: Insights From Cardiovascular Aging Science. JAMA Cardiology. 2020.

233. Yazdanpanah F, Hamblin MR, Rezaei N. The immune system and COVID-19: Friend or foe? Life Sci. 2020;256:117900.

234. Siripanthong B, Nazarian S, Muser D, Deo R, Santangeli P, Khanji MY, et al. Recognizing COVID-19-related myocarditis: The possible pathophysiology and proposed guideline for diagnosis and management. Heart rhythm. 2020:S1547-5271(20)30422-7.

235. Lippi G, South AM, Henry BM. Electrolyte imbalances in patients with severe coronavirus disease 2019 (COVID-19). Annals of Clinical Biochemistry. 2020;57(3):262–5.

236. Driggin E, Madhavan MV, Bikdeli B, Chuich T, Laracy J, Biondi-Zoccai G, et al. Cardiovascular Considerations for Patients, Health Care Workers, and Health Systems During the COVID-19 Pandemic. Journal of the American College of Cardiology. 2020;75(18):2352–71.

237. Lazzerini PE, Boutjdir M, Capecchi PL. COVID-19, arrhythmic risk and inflammation: mind the gap! Circulation. 2020.

238. Wu C-I, Postema PG, Arbelo E, Behr ER, Bezzina CR, Napolitano C, et al. SARS-CoV-2, COVID-19, and inherited arrhythmia syndromes. Heart rhythm. 2020:S1547-5271(20)30285-X.

239. Liu K, Fang Y-Y, Deng Y, Liu W, Wang M-F, Ma J-P, et al. Clinical characteristics of novel coronavirus cases in tertiary hospitals in Hubei Province. Chinese medical journal. 2020.

240. Tomasoni D, Italia L, Adamo M, Inciardi RM, Lombardi CM, Solomon SD, et al. COVID-19 and heart failure: from infection to inflammation and angiotensin II stimulation. Searching for evidence from a new disease. European Journal of Heart Failure.n/a(n/a).

241. Medina de Chazal H, Del Buono MG, Keyser-Marcus L, Ma L, Moeller FG, Berrocal D, et al. Stress Cardiomyopathy Diagnosis and Treatment. <span class=“subtitle”><em>JACC</em> State-of-the-Art Review</span>. 2018;72(16):1955–71.

242. Pasqualetto MC, Secco E, Nizzetto M, Scevola M, Altafini L, Cester A, et al. Stress Cardiomyopathy in COVID-19 Disease. European journal of case reports in internal medicine. 2020;7(6):001718-.

243. Roca E, Lombardi C, Campana M, Vivaldi O, Bigni B, Bertozzi B, et al. Takotsubo Syndrome Associated with COVID-19. European journal of case reports in internal medicine. 2020;7(5):001665-.

244. Giustino G, Croft LB, Oates CP, Rahman K, Lerakis S, Reddy VY, et al. Takotsubo Cardiomyopathy in Males with Covid-19. Journal of the American College of Cardiology. 2020:S0735-1097(20)35551-0.

245. Nguyen D, Nguyen T, De Bels D, Castro Rodriguez J. A case of Takotsubo cardiomyopathy with COVID 19. European heart journal cardiovascular Imaging. 2020:jeaa152.

246. Chadha S. ‘COVID-19 Pandemic’ Anxiety induced Tako-tsubo Cardiomyopathy. QJM: monthly journal of the Association of Physicians. 2020:hcaa135.

247. Fung G, Luo H, Qiu Y, Yang D, McManus B. Myocarditis. Circulation Research. 2016;118(3):496–514.

248. Hu H, Ma F, Wei X, Fang Y. Coronavirus fulminant myocarditis saved with glucocorticoid and human immunoglobulin. European heart journal. 2020:ehaa190.

249. Zeng JH, Liu Y-X, Yuan J, Wang F-X, Wu W-B, Li J-X, et al. First case of COVID-19 infection with fulminant myocarditis complication: case report and insights. 2020.

250. Corrales-Medina VF, Madjid M, Musher DM. Role of acute infection in triggering acute coronary syndromes. Lancet Infect Dis. 2010;10(2):83–92.

